# Cross-Ancestry Polygenic Risk Scores Enhance Alzheimer’s Disease Risk Prediction in Multiethnic Cohorts

**DOI:** 10.1101/2025.10.03.25337285

**Authors:** Meri Okorie, Caroline Jonson, Alexis P. Oddi, Patricia A. Castruita, Brian Fulton-Howard, Kristine Yaffe, Jennifer S. Yokoyama, Chinedu Udeh-Momoh, Shea J. Andrews, the Alzheimer’s Disease Sequencing Project and the Healthy Aging Brain Study - Health Disparities

**Affiliations:** UCSF School of Pharmacy and the UCSF School of Medicine, University of California, San Francisco, San Francisco, USA; Pharmaceutical Sciences and Pharmacogenomics Program, University of California, San Francisco, San Francisco, USA; Department of Neurology and Weill Institute for Neurosciences, University of California, San Francisco, San Francisco, USA; Memory and Aging Center, University of California, San Francisco, San Francisco, CA, USA; Department of Radiology and Biomedical Imaging, University of California, San Francisco, San Francisco, CA, USA; Department of Psychiatry and Behavioral Sciences, University of California, San Francisco, San Francisco, USA; Department of Epidemiology and Biostatistics, University of California, San Francisco, San Francisco, CA, USA; San Francisco Veterans Affairs Health Care System, San Francisco, CA, USA; The Center for Population Brain Health, University of California, San Francisco, San Francisco, CA, USA; Center for Alzheimer’s and Related Dementias, National Institutes of Health, Bethesda, MD, USA; Ronald M Loeb Center for Alzheimer’s disease, Icahn School of Medicine at Mount Sinai, New York, NY, USA; Department of Neuroscience, Icahn School of Medicine at Mount Sinai, New York, NY, USA; DataTecnica LLC, Washington, DC USA; Department of Epidemiology and Prevention, Wake Forest University School of Medicine, USA; Brain and Mind Institute, Aga Khan University, Nairobi, Kenya

**Keywords:** Alzheimer’s disease, polygenic risk score, AD-PRS, population genetics

## Abstract

**INTRODUCTION:** Genome-wide association studies (GWAS) have identified 80+ genetic loci associated with Alzheimer’s disease (AD), enabling the development of polygenic risk scores (PRS). However, the predictive accuracy of PRS in diverse populations remains low. Here, we evaluated the predictive accuracy of single-, multi-, and cross-ancestry AD-PRS models across multi-ancestral populations.

**METHODS:** We used AD GWAS summary statistics from European, African, Amerindian, and East Asian populations to construct AD-PRS for each target population. Model performance was assessed by estimating odds ratios, R^2^, and AUC.

**RESULTS:** The cross-ancestry Bayesian PRS model demonstrated the highest predictive performance in non-European populations. It was significantly associated with poorer cognitive function, lower Aβ_42_ CSF levels, and the most severe category of Aβ and tau neuropathological burden, as well as a clinical AD latent variable in a multi-ancestral validation cohort.

**DISCUSSION:** Inclusive genetic datasets and cross-ancestry PRS models are needed to enhance the transportability of AD-PRS across multi-ancestral populations.

**Research in context:**

1. Systematic review: Using diverse GWAS datasets to construct AD-PRS is a promising yet underexplored approach to improve risk prediction accuracy across different populations. Integrating diverse base GWAS datasets and evaluating various PRS models can enhance model performance, but such approaches have not yet been widely applied to multi-ancestral cohorts to measure AD risks or abnormalities in biomarkers.
2. Interpretation: Incorporation of ancestrally diverse base GWAS datasets enhanced the association between PRS and AD risk across multiple populations. Leveraging both these diverse discovery datasets and a Bayesian framework markedly improved model performance, extending its potential clinical applicability beyond AD case-control classification to the prediction of biomarker abnormalities.
3. Future directions: Future research should prioritize the validation of cross-ancestry PRS models in larger and more heterogeneous populations, alongside systematic benchmarking against an expanding repertoire of PRS methodologies. Clinical implementation of AD-PRS will require rigorous validation in large, diverse as well as community-based populations to ensure reproducibility and generalizability, thereby enhancing its translational relevance.

**Highlights:**

- Single-ancestry PRS is only predictive in participants of European populations.
- Cross-ancestry PRS improves risk predictions in non-European participants.
- Cross-ancestry PRS is associated with abnormal Aβ and tau pathology and cognitive decline
- Cross-ancestry PRS is associated with the AD latent variable in a multi-ancestral cohort

## BACKGROUND

The abundance of GWAS and the growing number of disease-associated genetic variants have enabled the development of polygenic risk scores (PRS). PRSs calculate an individual’s genetic liability for a given trait or a disease by aggregating all the variants weighted by their effect size estimates derived from GWAS summary statistics^1^. In some diseases, at-risk individuals identified through PRS exhibit up to 20-fold higher than the carrier frequency of rare monogenic mutations conferring comparable risk^2^. These findings highlight the potential of PRS to identify at-risk individuals who may not be captured through family history or monogenic variants, creating opportunities to modify risk factors and personalize care before clinical symptoms emerge^3^.

AD GWAS have greatly advanced our understanding of AD genetic architecture, with heritability estimated to be 40-60%^4–7^, and facilitate the development of AD-PRS. AD-PRS show good predictive accuracy, with the best-performing model having an area under the curve (AUC) of 0.74 while adjusting for *APOE* ε2 and ε4 status^8^. The predictive accuracy further increases to 81% when applied to neuropathologically confirmed cases and controls or when using extreme PRS cutoffs^9,10^. The high predictive accuracy of AD-PRS demonstrates its potential utility for risk assessment. However, clinical translation has been slow, partly due to inconsistent performance across studies that raises concerns about generalizability^11–15^.

A major driver of the lack of generalizability of AD-PRS stems from the lack of diversity in GWAS training datasets. Most AD-PRS models have been derived from European ancestry GWAS, given the scarcity of large studies in non-European or admixed populations^16,17^. As a result, AD-PRSs derived from European datasets show markedly reduced predictive accuracy when applied to non-European ancestry populations, with performance declining as genetic distance from the training population increases^16–18^. For instance, AD-PRS derived from African American GWAS datasets outperformed those based on European or multi-ancestry AD-PRS in the African cohort, with some variability across different regions^13^. Similarly, including variants identified specifically in African populations has been shown to improve the accuracy of PRS across ancestrally diverse and admixed populations^15^. In some cases, AD-PRS derived from European GWAS data remains associated with AD risk in non-European populations by careful tuning of pruning and thresholding (P+T) parameters^18^; however, such optimizations may not be practical for routine clinical use. These trends highlight the limited portability of European-derived PRS and the potential to exacerbate existing health disparities if such models are applied indiscriminately^16^.

The need to address these disparities is particularly urgent given the disproportionate burden of ADRD among older Black and Latinx individuals compared to non-Latinx White individuals^19^. Yet, relatively few studies have systematically evaluated the performance of different AD-PRS models across ancestrally diverse populations, largely due to the limited availability of training and validation datasets. By increasing GWAS sample sizes of historically underrepresented populations and matching the statistical power of European ancestry GWAS, the accuracy of PRS can improve^15,20–24^. Although current non-European AD GWAS are notably smaller than those of European ancestry, multi-ancestry PRS derived from smaller ancestry-matched GWAS have been shown to outperform PRS derived from larger European GWAS^25^. Alternatively, cross-ancestry PRS methods aim to jointly model GWAS summary statistics from multiple ancestry groups to improve PRS prediction within a specified target ancestry^22,23^.

The present study evaluates the predictive accuracy of AD-PRS computed using single-, multi-, and cross-ancestry PRS models in multi-ancestral populations. We expect that AD-PRS will exhibit reduced predictive accuracy in populations not represented in the training data. Beyond disease diagnosis, we also assessed the clinical validity of PRS by examining whether the best-performing model was associated with AD endophenotypes, including biomarkers and cognitive domains within the A/T/N framework^26,27^. This evaluation provides insight into whether PRS can capture early disease-related processes, complementing risk stratification for AD.

## METHODS

### 2.1 Datasets and Quality Control (QC)

#### 2.1.1 Genome-Wide Association Study (GWAS) Datasets

We utilized summary statistics from the largest publicly available ancestry-specific GWAS datasets on AD and related dementias (ADRD) to date, including participants of European (Bellenguez et al.^5^: 39,106 cases, 46,828 proxy cases, 401,577 controls; FinnGen Release 6: 7,329 cases, 131,102 controls), African (Kunkle et al.: 2,748 cases, 5,222 controls^28^), East Asian (Shigemizu et al.^29^: 3,962 cases, 4,074 controls), and Caribbean Hispanic descent (Columbia University Study: 1,088 cases, 1,152 controls). We also utilized multi-ancestry meta-analysis (MAMA) GWAS summary statistics, combining data from the five ancestry-specific studies listed above (Lake et al.^30^: 54,233 AD cases, 46,828 proxy cases, 543,127 controls) to construct a multi-ancestry PRS.

#### 2.1.2 Alzheimer’s Disease and Sequencing Project

Data from the Alzheimer’s Disease Sequencing Project (ADSP) Release 4 whole genome sequencing dataset were accessed from the National Institute on Aging Genetics of Alzheimer’s Disease Data Storage Site (NIAGADS) (https://dss.niagads.org/). The participants in the ADSP are recruited from large, existing family-based and case/control studies that provide comprehensive phenotypic and genetic data. The ADSP has made a concerted effort to include participants from multi-racial and -ethnic groups, as many earlier genetic studies have predominantly involved individuals of European descent. AD diagnosis was harmonized across the cohorts by the ADSP Phenotype Harmonization Consortium (ADSP-PHC)^31^. The ADSP-PHC also harmonized AD endophenotype measurements, including cognitive function (language, memory, executive functions; total N = 11,046), cerebrospinal fluid (CSF) biomarkers (Tau, pTau_181_, Aβ_42_; total N = 815), and neuropathology (CERAD, N = 2,727; Thal Phase, N = 1,186; BRAAK staging, N = 2,725)^32,33^. For exclusion, A total of 515 participants diagnosed with mild cognitive impairment (MCI) were excluded from the analysis. An additional 117 participants from a family-based study that overlapped with case/control studies were removed. Additionally, 488 samples were removed due to missing genotype data, and 5,763 participants were removed due to missing AD diagnosis information.

A standardized Quality Control (QC) pipeline was applied to the WGS datasets. Variants failing to meet predefined quality thresholds were excluded, including those with a minimum depth of coverage below 10, genotype quality below 20, and minor allele frequency (MAF) below 0.01. Variants with missing rsIDs were assigned a chromosome position ID. To further assess quality, sample-level and variant-level QC were performed using GenoTools^34^ to undergo a series of QC checks. Sample-level QC includes call rate (≥ 0.95), sex concordance (male: F ≥ 0.99, female: F ≥ 0.03), and relatedness between samples (cutoff = 0.0884). Variant-level QC was performed to examine non-random missingness by haplotypes, Hardy-Weinberg equilibrium (≤ 1e-4), and genotype missingness (< 0.05). 2,743 participants were removed due to sex discrepancy, 359 participants were removed due to heterozygosity pruning, and 3,164 participants were removed due to the relatedness check. The African GWAS summary statistics were derived from participants in the Alzheimer’s Disease Genetic Consortium (ADGC), which includes sample overlaps with those from the ADSP. To retain sample independence between the base and the target datasets and avoid PRS inflation caused by sample overlap^35^, relationship inference was performed using software, KING^36^, to identify and exclude samples based on relatedness. Pairwise samples with kinship coefficients ≥ 0.354^36^, 2,776 participants, were removed from the ADSP target datasets. After the main GenoTools filters and excluding participants with missing data, 19,398 participants (7,111 cases and 12,287 controls) remained for the downstream analysis.

Genetic ancestry was determined using a principal component analysis (PCA) with the 1000 Genome Project (1KG) + Human Genome Diversity Project (HGDP) as the reference panel, followed by classification via a random forest algorithm implemented in pgsc_calc^37^. Out of 19,398 participants, 8,043 (41.5%) participants were classified as European ancestry, 1,945 (10.0%) as African ancestry, 6.901 (35.6%) as Amerindian ancestry, 2,365 (12.2%) as Central/South Asian, 72 (0.37%) participants were assigned East Asian ancestry, and 72 (0.37%) participants were assigned as Middle Eastern ancestry. Participants of East Asian and Middle Eastern ancestries were removed due to small sample size, while participants of Central South Asian ancestry were excluded due to imbalanced case-control ratios (18 cases to 2,347 controls). Global genetic admixture for each sample was estimated using ADMIXTURE version 1.3^38^. Briefly, we identified a representative reference panel composed of samples distributed by the 1KG. The reference panel and target ADSP dataset were subset to include only overlapping SNPs. We performed an unsupervised admixture analysis of the reference dataset, setting K = 5. The target samples were then projected onto the population structure learned from the reference panel.

### 2.2 Healthy Brain Aging Study - Health Disparities (HABS-HD)

#### 2.2.1 Participants

The Health & Aging Brain Study - Health Disparities (HABS-HD) cohort comprises participants from Black, Mexican American, and non-Hispanic White populations recruited at the University of North Texas Health Science Center, Fort Worth, Texas, USA^39,40^. The study collected information on demographics, AD biomarkers, neuroimaging, clinical history, and genomics, providing a comprehensive resource for AD and related dementia research. Eligibility to participate in the study was restricted to generally healthy individuals without serious mental or medical conditions, who were willing to undergo sample collections and neuroimaging procedures. We utilized participant data from the HABS-HD release 5 baseline visit 1. We further applied the exclusion criteria to remove participants younger than 55 years old, those without genetic data, and those with plasma biomarker values identified as outliers. Outliers were detected following the published methodology using the interquartile range approach^41^. All HABS-HD participants (and/or their legal guardians) signed written informed consent to participate in the study.

#### 2.2.2 AD endophenotypes

HABS-HD participants have undergone extensive phenotyping for AD endophenotypes, including plasma A/T/N biomarkers, brain morphometry, amyloid and tau PET, and neuropsychological testing, as previously described^40^. CSF and For downstream analyses of each endophenotype, we applied the following approaches:

##### Plasma biomarkers

Aβ_40_, Aβ_42_, pTau_181_, Tau, and NfL values were natural log-transformed to reduce skewness, and then standardized to z-scores (mean = 0, SD = 1) across the study samples. *Brain morphomtry:* Cortical thickness was defined as the surface area-weighted average of cortical thickness in the right and left entorhinal cortex, fusiform, and inferior and middle temporal cortex. Hippocampal volumes was determined by taking the mean volume of the right and left hippocampal volume determined on a T1-weighted volume scan. *neuropsychological testing:* Composite scores for each cognitive domain (memory, language, and executive function) were constructed by averaging z-score-normalized test scores from the neuropsychological battery. The memory domain was assessed using immediate and delayed recall from the Wechsler Memory Scale (WMS-III) Logical Memory and the Spanish English Verbal Learning Test (SEVLT). The language domain was assessed using Letter Fluency and

##### Animal Naming tests

The executive function domain was assessed using the WMS-III Digit Span and the Trail Making Test, Parts A and B. In addition, the Mini-Mental State Examination (MMSE) and Clinical Dementia Rating (CDR) scale were administered to all participants as part of the neuropsychological assessment. *Amyloid PET:* For amyloid positron emission tomography (PET) scan variables, the global standardized uptake value ratio (SUVR) was derived by normalizing tracer uptake to the whole cerebellum. Amyloid PET positivity was defined using a SUVR threshold of 1.08.

#### 2.2.3 Genome-wide Genotyping and QC

Participants were genotyped on the Illumina Global Screening Array (GSA) and genotype data underwent stringent quality control (QC) checks using an in-house Snakemake Pipeline^42^. Variants were excluded if the call rate was <0.95 and not in Hardy–Weinberg equilibrium (p < 1 × 10–6), and samples excluded if the call rate was <0.95, discordant sex was reported based on X chromosome heterozygosity, excessive/insufficient heterozygosity, and cryptic relatedness. Related individuals were determined within and across cohorts by identity-by-descent (IBD) using KING^36^, with individuals excluded based on a proportion of IBD <0.1875, corresponding to less than halfway between second- and third-degree relatives. Single-nucleotide polymorphisms that were not directly genotyped were imputed on the TOPMed Imputation Server^43^. Ancestry groups were imputed separately using all ethnicities of the TOPMed reference panel (Version R3), with Eagle used for phasing and Minimac3 used for imputation. Following imputation, poorly imputed (r2 < 0.3) or rare (MAF < 0.01) variants were removed, and the ancestry groups were merged for joint analysis. Following this merger, variants with low call rate due to differential imputation (<95%) were removed, and then samples with low call rates (<95%) were removed. The final dataset contained 2,559 samples with ∼22M high-quality SNPs per individual.

Of the total of 2,559 participants that were included in the downstream statistical analysis, 1,040 were of European ancestry, 891 were of Amerindian ancestry, and 574 were of African ancestry based on genetic ancestry inference (Refer to ancestry assignment in 2.3.2).

### 2.3 PRS Construction

#### 2.3.1 PRS Scorefile Generation

PRSice was used to implement P+T to construct four AD-PRS, stratified by EUR ancestry and MAMA ancestry, at two p-value thresholds. P+T generates scorefiles for SNPs that exceed a given p-value threshold (P_T_) and its corresponding effect size estimated from an independent GWAS. Scorefiles for the single-(EUR) and multi-ancestry (MAMA) PRS models were constructed using SNPs with a p-value < 0.1 (P_T0.1_) and genome-wide significance (GWS) (P_TGWS_). SNPs from apolipoprotein E (*APOE)* regions ± 500kb (GRCh38, chr19:bp44405791-45409393) were excluded from all the base datasets. A p-value threshold of 0.1 in the construction of PRS has been previously shown to have the best predictive accuracy and will be tested in this study^8^. Linkage disequilibrium (LD) clumping using 1KG + HGDP LD reference panels, with an r^2^ threshold of 0.1 and clumping distance of 250kb, was used to select independent SNPs. For the single-ancestry PRS model, the 1KG + HGDP reference panel was filtered for European-only samples, and the whole 1KG + HGDP reference panel was used for the multi-ancestry PRS model.

PRS-CSx was used to construct a scorefile for the cross-ancestry AD-PRS model, a method that improves cross-population polygenic prediction by jointly modeling GWAS summary statistics from multiple populations^23^. The global shrinkage parameter (phi) was set to “auto”, which allows for automatic estimation of the effect size from the datasets. Additionally, the “meta” parameter was set to ‘true’ to combine the SNP effect sizes across populations using an inverse-variance-weighted meta-analysis. The Hapmap SNP set was used to filter SNPs from the target dataset, and the summary statistics for ancestry-specific AD GWAS were used to construct the scorefile. SNPs from *APOE* regions ± 500kb (GRCh37, chr19:bp44909053-45912650) were excluded from the final scorefile prior to PRS calculation. Liftover was automatically applied during PRS calculation to concert scorefile build to GRCh38. The same methodology was applied to the HABS-HD cohort with the same parameters and base GWAS datasets.

#### 2.3.2 PRS Calculation and Ancestry Normalization

To mitigate ancestry-related confounding due to differences in PRS distribution and to enable more accurate PRS comparisons across ancestrally diverse populations^44^, we used pgsc_calc^37^ to construct PRS and normalize for ancestry differences in the ADSP and the HABS-HD cohort using the PCA method^45^. PCA was performed on the reference panel to derive principal components (PCs) representing global genetic variation. Target individuals were then projected onto these PCs to determine their position in genetic ancestry space. A RandomForest classifier, trained on the reference panel’s PCA loadings (default: 10 PCs), was used to predict the reference population to which each target sample was most similar^46^. Following PC projection and ancestry assignment, polygenic risk scores (PRS) were normalized by ancestry. First, a regression model predicted PRS from PC loadings, and the residuals were used to center the PRS distribution at zero across ancestry groups (znorm1). A second regression on the squared residuals adjusted for variance, standardizing the PRS spread to approximately one (znorm2). All five PRS models underwent ancestry normalization, and znorm2 scores were used for the downstream analysis (Supplementary Fig. 4).

### 2.5 Statistical Analysis

#### 2.5.1 AD Risk

Logistic regression models were used to evaluate the associations of EUR, MAMA, and PRS-CSx PRS models with AD, adjusting for age at baseline, sex, *APOE* genotypes, and 10 PCs. The *APOE* status was encoded as a categorical variable, categorized as *APOE* ε2+ carriers (ε2/ε2, ε2/ε3) and ε4+ carriers (ε2/ε4, ε3/ε4, or ε4/ε4), with the ε3/ε3 allele serving as the reference group. Model performance was evaluated via Area Under the Receiver Operating Curve (AUCROC) to assess model discrimination. R2redux^47,48^ was used to calculate and statistically compare the differences in R^2^ for the AD outcome between models. For P+T PRS models, only the GWS thresholds were included and compared to each other and to PRS-CSx to test disease-relevant SNPs (P+T) vs leveraging all SNPs (PRS-CSx). Calibration of AD-PRS was assessed to evaluate the estimated probability of AD against the observed probability using PRS as a predictor.

#### 2.5.2 Endophenotypes

In the ADSP, multiple linear regression models were fit to assess the relationship between PRS-CSx and (i) cerebrospinal fluid biomarkers (i.e., pTau_181_, total tau, and Aβ_42_), and (ii) cognitive domain scores (i.e., language, memory, and executive function). Models were adjusted for age, sex, *APOE* genotype, PC1-10, and study cohort. Models for cognitive outcomes were additionally adjusted for years of education. Ordinal logistic regression models, performed using the polr function in the MASS R package, were fitted for Thal phase, CERAD score, and Braak staging as ordinal outcomes, adjusting for age, sex, *APOE* genotype, PC1-10, and study cohort. All p-values were adjusted for multiple testing at 5% false discovery rate (FDR). Genetic ancestry stratification was performed for cognitive outcomes. CSF biomarkers and other cognitive measures had insufficient sample sizes for ancestry-stratified analyses.

Similarly in the HABS-HD, multiple linear regression models were fit to assess the associations between PRS-CSx and (i) plasma biomarkers (i.e., Aβ_40_, Aβ_42_, pTau_181_, Tau, and NfL), (ii) neuroimaging variables (i.e., cortical thickness and hippocampal volume), (iii) composite scores for each domain (i.e., language, memory, and executive function), and (iv) global cognition tests (i.e., CDR and MMSE). Models were adjusted for age, sex, *APOE* genotype, and PC1-4. Additionally, BMI and eGFR were accounted for the plasma biomarkers. Intracranial volume was accounted for the neuroimaging variables. Years of education were accounted for the neuropsychological test battery variables. Both raw and FDR-adjusted p-values (5%) were reported.

#### 2.5.3 AD latent variable

To validate AD-PRS generated by PRS-CSx in an external cohort, we used structural equation modeling to construct an AD latent variable, indicated by Aβ and tau pathology in HABS-HD. 2,297 participants were included in the structural equation modeling (SEM) analysis using the R package lavaan. Confirmatory factor analysis was used to derive an AD latent variable, indicated by plasma pTau_181_, Aβ PET positivity, and Aβ PET global SUVR. Prior to model fitting, we applied a two-stage least squared approach using instrumental variables. Aβ PET measures were adjusted for MRI scanner type, and plasma pTau_181_ was adjusted for body mass index and estimated glomerular filtration rate to account for the distribution and clearance of the plasma biomarker. Using SEM, the AD latent variable was regressed on AD-PRS, adjusting for age, sex, *APOE* ε4+ status, and population stratification using genetic principal components (4 PCs). Multigroup analysis was also performed to measure the associations between AD-PRS and AD latent variable across ancestry groups. Full information maximum likelihood (FIML) estimation was used to estimate model parameters. Model fit was assessed using the Comparative Fit Index (CFI), Tucker-Lewis Index (TLI), Root Mean Square Error of Approximation (RMSEA), and Standardized Root Mean Square Residual (SRMR). Thresholds of CFI and TLI ≥ 0.95, and RMSEA and SRMR ≤ 0.06, were used to indicate an excellent model fit^49,50^.

All analyses were conducted using R version 4.4.3.

## RESULTS

### 3.1 Patient characteristics

After exclusion, we retained 19,398 participants from ADSP, including 12,287 classified as cognitively normal and 7,111 diagnosed with AD (Table 1). The cohort had a mean age of 68 ± 10 years and consisted of 60% females. European ancestry was the most common (41%), followed by Amerindian (36%) and African ancestries (10%), respectively. The majority of people with AD were *APOE* ε4 allele carriers. A similar profile is presented in the HABS-HD, with 61% female with a mean age of 69 ± 8 years. The cohort primarily consists of participants of European (41%), Amerindian (35%), and African ancestry (22%), with the majority of people with AD carrying at least one *APOE* ε4 allele (Table 2).

**Table 1.** Characteristics of the ADSP cohort, including age, sex, genetic ancestry, and *APOE* genotype.

MAIN FIGURES
| Characteristics |  |  |  |
| --- | --- | --- | --- |
| Diagnosis |  | CN | AD |
| **Demographics** | 19,398 | 12,287* | 7,111* |
| Age |  | 68 (10) | 73 (9) |
| Sex |  |  |  |
| Male |  | 4,782 (39%) | 2,928 (41%) |
| Female |  | 7,505 (61%) | 4,183 (59%) |
| Ancestry |  |  |  |
| European |  | 3,637 (30%) | 4,406 (62%) |
| African |  | 1,422 (12%) | 523 (7.4%) |
| Amerindian |  | 4,818 (39%) | 2,083 (29%) |
| Central South Asian |  | 2,347 (19%) | 18 (0.3%) |
| East Asian |  | 32 (0.3%) | 40 (0.6%) |
| Middle Eastern |  | 31 (0.3%) | 41 (0.6%) |
| *APOE* Genotype |  |  |  |
| e3/e3 |  | 8,065 (66%) | 3,007 (42%) |
| e2+ |  | 1,171 (9.5%) | 315 (4.4%) |
| e4+ |  | 3,051 (25%) | 3,789 (53%) |
| **CSF** | 621 | 325 | 296 |
| Aβ<sub>42</sub> | | 0.33 (0.97) | -0.63 (0.86) |
| pTau |  | -0.24 (0.85) | 0.63 (0.93) |
| Tau |  | -0.25 (0.89) | 0.57 (0.91) |
| **Cognitive Domains** | 8434 | 4,446 | 3,988 |
| Memory |  | 0.65 (0.53) | -0.60 (0.78) |
| Language |  | 0.39 (0.71) | -0.33 (0.79) |
| Executive Function |  | 0.41 (0.83) | -0.43 (0.89) |
| **Thal Phase** | 885 | 167 | 718 |
| None |  | 47 (28%) | 22 (3%) |
| Phase 1 |  | 33 (19%) | 10 (1%) |
| Phase 2 |  | 18 (11%) | 17 (2%) |
| Phase 3 |  | 33 (20%) | 54 (8%) |
| Phase 4 |  | 26 (16%) | 127 (18%) |
| Phase 5 |  | 10 (6%) | 488 (68%) |
| **CERAD Score** | 2,337 | 807 | 1,629 |
| None |  | 407 (58%) | 55 (3%) |
| Sparse/Possible |  | 185 (26%) | 79 (5%) |
| Moderate/Probable |  | 82 (12%) | 361 (22%) |
| Definite/Frequent |  | 34 (5%) | 1134 (70%) |
| **BRAAK Staging** | 2,337 | 707 | 1,630 |
| None |  | 34 (5%) | 10 (0.6%) |
| BRAAK Stage I |  | 122 (17%) | 17 (1%) |
| BRAAK Stage II |  | 187 (26%) | 41 (3%) |
| BRAAK Stage III |  | 195 (28%) | 90 (6%) |
| BRAAK Stage IV |  | 139 (20%) | 222 (14%) |
| BRAAK Stage V |  | 26 (4%) | 494 (30%) |
| BRAAK Stage VI |  | 4 (0.6%) | 756 (46%) |
| *Mean (SD); n (%) |  |  |  |

**Table 2.** Characteristics of the HABS-HD cohort, including age, sex, genetic ancestry, and *APOE* genotype.

| Characteristics |  |  |  |
| --- | --- | --- | --- |
| Diagnosis |  | CN | MCI + Dementia |
| **Demographics** | 2,559 | 1,855* | 704* |
| Age |  | 67 (8) | 71 (8) |
| Sex |  |  |  |
| Female |  | 1,549 (61%) | 16 (52%) |
| Male |  | 979 (39%) | 15 (48%) |
| Ancestry |  |  |  |
| European |  | 1,024 (41%) | 16 (52%) |
| Amerindian |  | 888 (36%) | 3 (9.7%) |
| African |  | 562 (23%) | 12 (39%) |
| Unknown |  | 54 | 0 |
| *APOE* Genotype |  |  |  |
| e3/e3 |  | 1,331 (63%) | 5 (26%) |
| e2+ |  | 228 (11%) | 2 (11%) |
| e4+ |  | 567 (27%) | 12 (63%) |
| Unknown |  | 402 | 12 |
| **Plasma** |  |  |  |
| Aβ<sub>42</sub>/Aβ<sub>40</sub> | | 0.02 (0.99) | -0.02 (1.02) |
| pTau |  | -0.10(0.90) | 0.26 (1.19) |
| Tau |  | -0.02 (-.97) | 0.07 (1.08) |
| **Neuroimaging** |  |  |  |
| Cortical Thickness |  | 2.75 (0.13) | 2.69 (0.17) |
| Hippocampal Volume |  | 0.12 (0.92) | -0.32 (1.13) |
| **PET** |  |  |  |
| Aβ SUVR | | 1.04 (0.15) | 1.37 (0.24) |
| Aβ Positivity | | 195 (19%) | 18 (90%) |
| **Cognitive Domains** |  |  |  |
| Language |  | 0.24 (0.75) | -0.62 (0.84) |
| Memory |  | 0.33 (0.67) | -0.85 (0.78) |
| Executive Function |  | -0.05 (0.48) | 0.12 (0.72) |
| **Global Cognition** |  |  |  |
| MMSE |  | 28.20 (2.11) | 25.37 (4.33) |
| CDR |  | 0.00 (0.00) | 2.07 (2.17) |
| *Mean (SD); n (%) |  |  |  |

The PCA projection of the ADSP revealed a high admixture profile among participants of Amerindian ancestry. Most of these participants clustered into two distinct patterns, separated from the European ancestry group, while some overlapped with the African ancestry group (Fig. 1B). Consistent with this, the admixture analysis showed that participants of Amerindian ancestry exhibited genetic heterogeneity, with contributions from European, Amerindian, and African ancestries (Fig. 1C).

**Figure 1.**
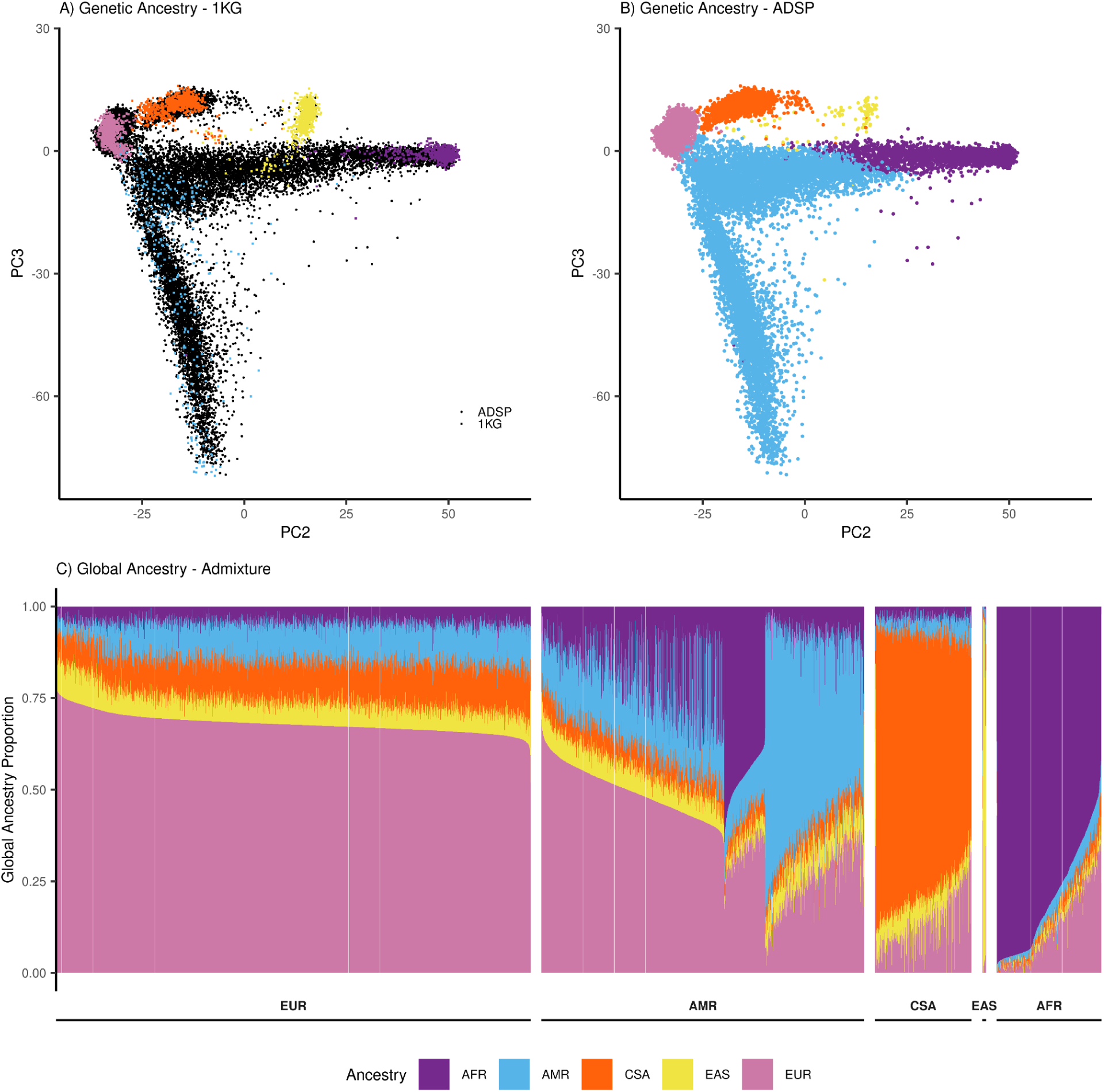
Genetic ancestry and global admixture analysis in the ADSP cohort. (A) Principal component analysis (PCA) of genetic ancestry using the 1000 Genomes (1KG) reference panel. ADSP participants (black dots) are overlaid onto 1KG reference populations (square dots). (B) PCA of ADSP participants only. (C) Global ancestry proportions estimated using admixture analysis, illustrating varying degrees of admixture across ancestry groups.

### 3.2 EUR Single-Ancestry PRS

The European-only, single-ancestry PRS model showed robust associations with AD risk in individuals of European ancestry at both PT_gws_ (OR [95%CI] = 1.44 [1.37-1.52], p = 5.53 x 10^−42^) and PT_0.1_ (OR [95%CI] = 1.38 [1.31-1.47], p = 1.69 × 10^−27^) (Fig. 2). Predictive performance also declined as the genetic distance for participants of African and Amerindian ancestry increased from the participants of European ancestry (R² = 0.013, 0.018, 0.026, respectively), and AUCROC metrics remained stable across models (Fig. 3; Supplementary Fig. 2). Association of EUR PRS remained statistically significant in the participants of Amerindian ancestry at both PT_gws_ (OR [95%CI] = 1.17 [1.10-1.24], p = 5.59 x 10^−7^) and PT_0.1_ (OR [95%CI] = 1.14 [1.06-1.21], p = 1.69 x 10^−4^), though the effect size was smaller. It showed no association with AD risk in participants of African ancestry at both PT_gws_ (OR [95%CI] = 1.08 [0.96-1.21], p = 0.22) and PT_0.1_ threshold (OR [95%CI] = 0.93 [0.81-1.04], p = 0.22).

**Figure 2.**
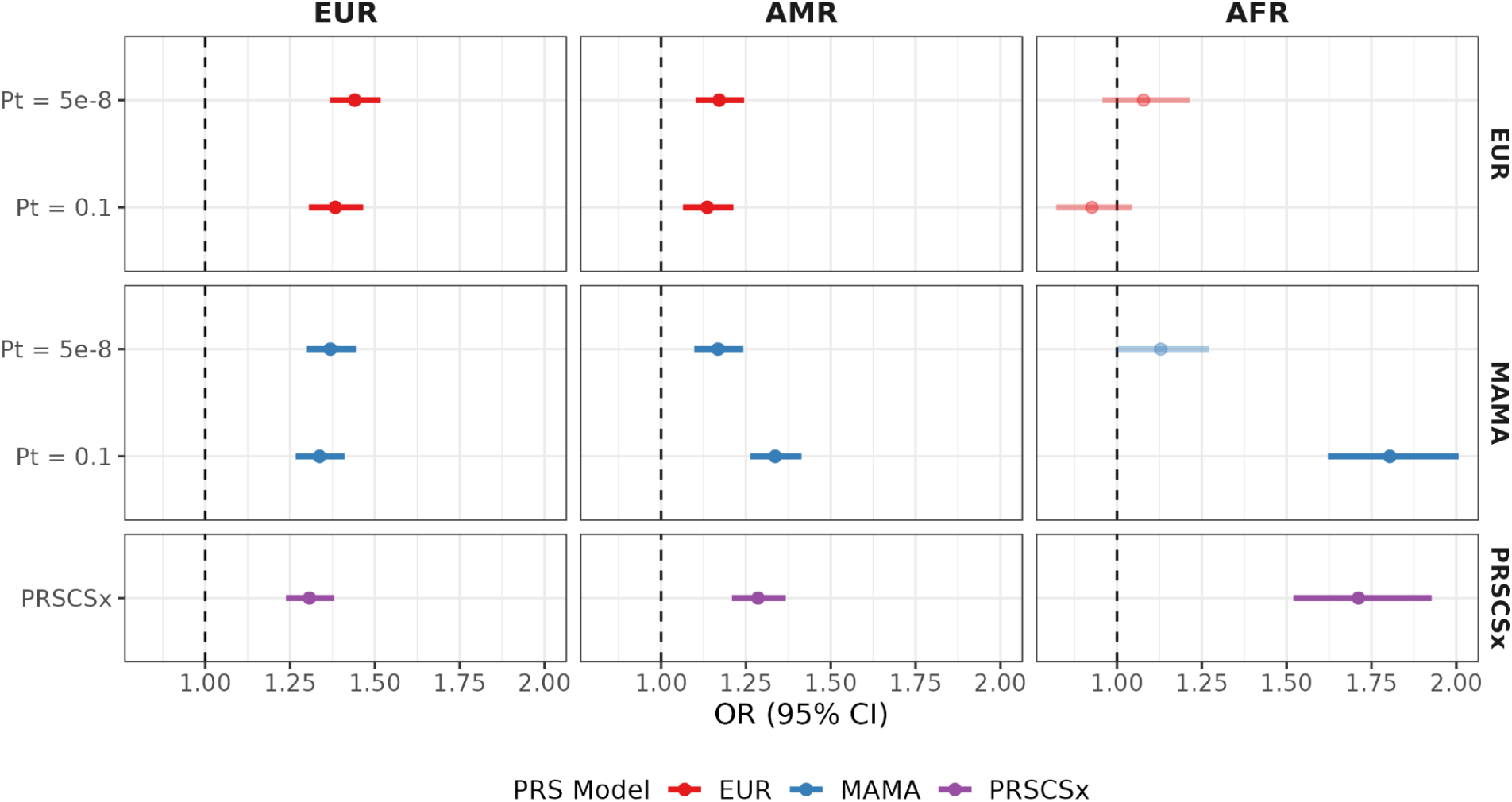
Odds ratio of ancestry-normalized single-(EUR), multi-(MAMA), and cross-ancestry (PRS-CSx) PRS models (right x-axis) stratified by genetic ancestry (top y-axis). P-values were adjusted for multiple testing using the Benjamini-Hochberg false discovery rate procedure, controlling the false discovery rate (FDR) at 5%.

**Figure 3.**
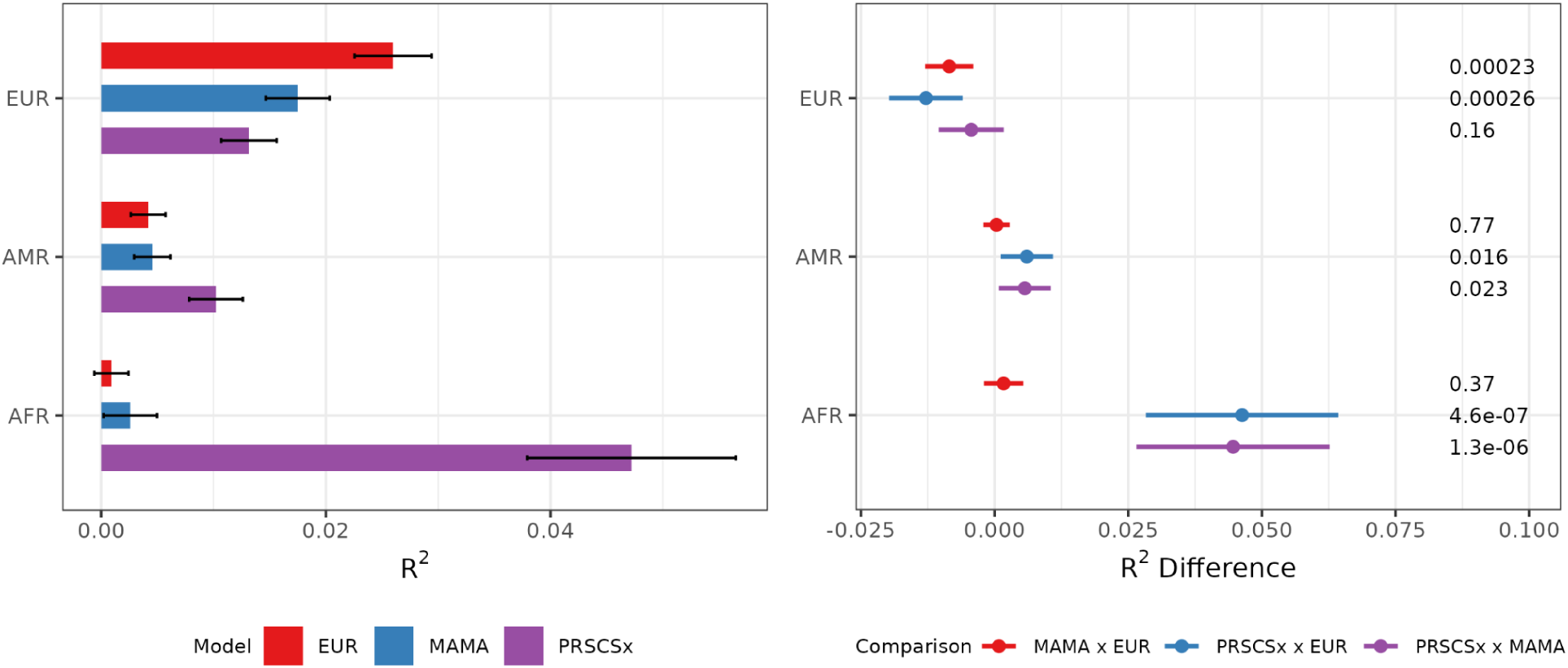
Comparison of model performance across ancestry groups. (Left) R^2^ for EUR PRS (GWS), MAMA PRS (GWS), and PRS-CSx PRS models in the participants of European (EUR), Amerindian (AMR), and African (AFR) ancestry groups. Error bars represent 95% confidence intervals. (Right) Differences in R^2^ between EUR versus MAMA PRS, EUR versus PRS-CSx PRS, and MAMA versus PRS-CSx PRS models. Error bars represent 95% confidence intervals of the difference.

### 3.3 MAMA Multi-Ancestry PRS

When leveraging the MAMA GWAS summary statistics, multi-ancestry PRS performance improved substantially in the participants of African and Amerindian ancestry while remaining comparable in the participants of European ancestry. In the participants of European ancestry, the MAMA PRS showed a similar association as EUR PRS with AD risk for PT_gws_ (OR [95%CI] = 1.44 [1.37-1.52], p = 5.53 × 10^−42^) and PT_0.1_ (OR [95%CI] = 1.38 [1.31-1.47], p = 1.69 × 10^−27^) (Fig. 2). PT_0.1_ PRS model showed a stronger association with AD risk than PT_gws_ PRS model in the participants of Amerindian ancestry (PT_gws_ (OR [95%CI] = 1.17 [1.10-1.24], p = 1.27 × 10^−6^) & PT_0.1_ (OR [95%CI] = 1.34 [1.26-1.41], p = 1.71 × 10^−23^). The effect was even larger in the participants of African ancestry with PT_0.1_ MAMA PRS model demonstrating 80% increase in the odds of developing AD while PT_gws_ remained nonsignificant (PT_gws_ (OR [95%CI] = 1.13 [1.00-1.27], p = 0.053) & PT_0.1_ (OR [95%CI] = 1.80 [1.62-2.01], p = 7.30 × 10^−27^) (Fig. 2).

### 3.4 PRS-CSx Cross-ancestry PRS

The PRS-CSx cross-ancestry model showed similar trends as PT_0,1_ MAMA PRS models in regards to the association with AD. It was associated with the highest AD risk in the participants of African ancestry (OR [95%CI] = 1.71 [1.52-1.93], p = 1.45 x 10^−18^), while remaining significant in the participants of Amerindian and European ancestry (OR [95%CI] = 1.29 [1.21-1.37], p = 1.76 x 10^−15^ & OR [95%CI] = 1.31 [1.24-1.38], p = 7.86 x 10^−22^, respectively) (Fig. 2). The model also achieved the highest R^2^ values in the participants of African and Amerindian ancestry (R^2^ = 0.047, 0.010, respectively) compared to other models (Fig. 3). Differences in R^2^ was also significant between PRS-CSx and EUR or MAMA PRS models in the participants of African ancestry (ΔR^2^ p = 4.6 x 10^−7^ & 1.3 x 10^−6^, respectively), showcasing PRS-CSx strong predictive performance. A less prominent yet similar trend was seen in the participants of Amerindian ancestry (ΔR^2^ p = 0.016 & 0.023, EURxPRS-CSx & MAMAxPRS-CSx).

Calibration analyses further illustrate that while all models calibrate best in the participants of European ancestry, and maintain high accuracy in the participants of Amerindian ancestry, predictive accuracy decays progressively with increasing genetic distance from Europeans, with the greatest calibration gains in African ancestry arising from ancestry-normalized MAMA and PRS-CSx PRS models (Supplementary Fig. 3, Supplementary Fig. 5).

### 3.5 AD endophenotypes

To measure PRS performance beyond their established utility in predicting AD risk, we further evaluated the association of the best-predictive PRS model, PRS-CSx, with key cognitive domains (memory, language, and executive function) and CSF biomarkers (Tau, pTau_181_, Aβ_42_). PRS-CSx demonstrated strong associations with poorer cognitive performance across multiple domains (β [95% CI]: Memory = 0.94 [0.93-0.95], p = 6.69 x 10^−18^, R² = 0.25, Language = 0.95 [0.94-0.97], p = 1.10 x 10^−12^, R² = 0.22, Executive function = 0.95 [0.94-0.97], p = 1.18 x 10^−9^, R² = 0.30) in the total cohort.

PRS-CSx demonstrated significant associations with poorer cognitive performance across multiple domains in participants of European ancestry (Memory (β [95% CI] = −0.08 [−0.099, −0.062], p = 3.71 x 10^−17^, R² = 0.26); Language (β [95% CI] = −0.059 [−0.077, −0.042], p = 2.87 x 10^−10^, R² = 0.11); and Executive function (β [95% CI] = −0.056 [−0.075, −0.037], p = 1.22 x 10^−8^, R² = 0.19). No associations were observed in participants of Amerindian and African ancestry across various cognitive domains. In African ancestry participants, language and executive function were not associated with PRS-CSx, with relatively wide confidence intervals due to small sample size, while memory showed no association in Amerindian ancestry participants. However, R² values were still substantial, ranging from 0.29–0.40 in African ancestry and 0.29–0.34 in Amerindian ancestry participants throughout the domains (Supplementary table 3). Similarly, PRS-CSx was associated with lower Aβ_42_ CSF levels (β [95% CI] = 0.91 [0.85-0.98], p = 0.015) (Fig. 4), but was not associated with Tau or pTau_181_. Furthermore, we evaluated the predicted probabilities of PRS-CSx across three neuropathological measures: the Consortium to Establish a Registry for Alzheimer’s Disease (CERAD) score, Thal Phase, and Braak Staging. Across all three measures, higher PRS-CSx values were associated with an increased probability of falling into the most severe category (e.g., “Frequent/Definite” for CERAD, “Phase 5” for Thal, and “Stage VI” for Braak), suggesting a strong relationship between PRS-CSx and advanced AD pathology (Fig. 4).

**Figure 4.**
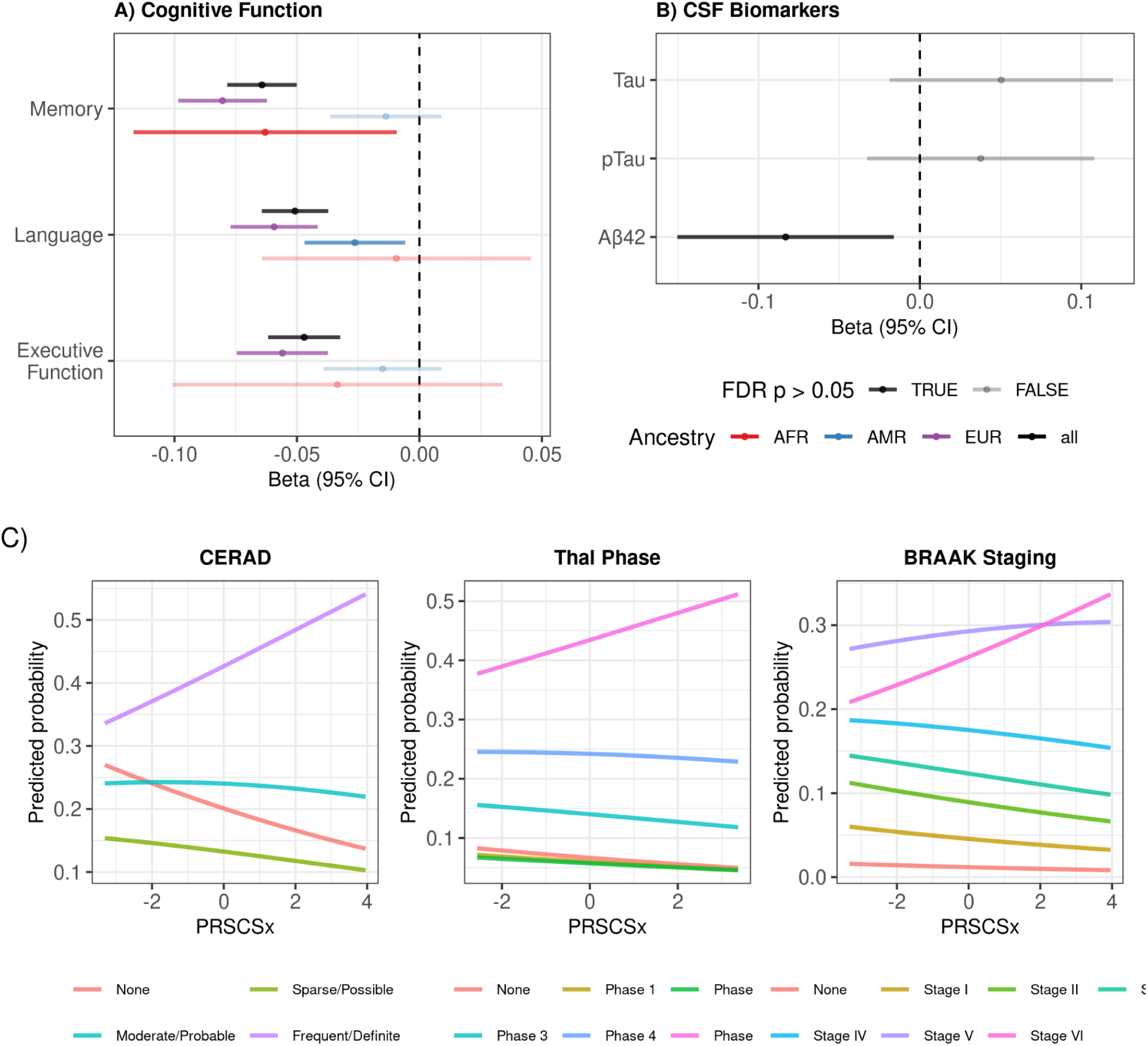
Associations of the best-predictive PRS model, cross-ancestry PRS-CSx, with A) cognitive function and B) cerebrospinal fluid (CSF) biomarkers in the ADSP. The dark blue color represents statistical significance after correction at a 5% false discovery rate (FDR). Cognitive function: N = 10,903 (AFR: 409, AMR: 2,888, EUR: 7,606), CSF biomarkers: N = 808. C) Predicted probabilities of neuropathological staging outcomes by PRS-CSx. CERAD: N = 2,713, Thal Phase: N = 1,175, BRAAK staging: N = 2,711. The sample sizes reported in Table 1 vary, as certain participants contributing to the endophenotype analyses lack an AD diagnosis.

### 3.6 AD latent variable

We further validated the association of the cross-ancestry AD-PRS in HABS-HD using AD endophenotypes. Furthermore, we constructed an AD latent variable based on pTau_181_ and Aβ biomarkers using SEM to more accurately model AD pathology. The AD-PRS cross-ancestry model was significantly associated with plasma Aβ_42_ and pTau_181_ levels in the total population. The association with pTau_181_ was also significant in the participants of European ancestry, but there were no other ancestry-specific effects in the participants of African and Amerindian ancestries. After FDR correction at 5%, there were no significant associations between PRS-CSx and all the endophenotypes (i.e., plasma biomarkers, brain morphometry, and cognition) (Supplementary Table 2).

The association between PRS-CSx and AD latent variable achieved statistical significance in the total cohort (Fig. 5), indicating that higher PRS was associated with greater AD-related pathology. The SEM model demonstrated strong fit, with CFI = 0.983, TLI = 0.963, RMSEA = 0.037, and SRMR = 0.021. However, the multigroup analysis of SEM analysis did not yield statistically significant associations between PRS-CSx and AD latent variables in the participants of Amerindian and African ancestry (Supplementary fig. 6).

**Figure 5.**
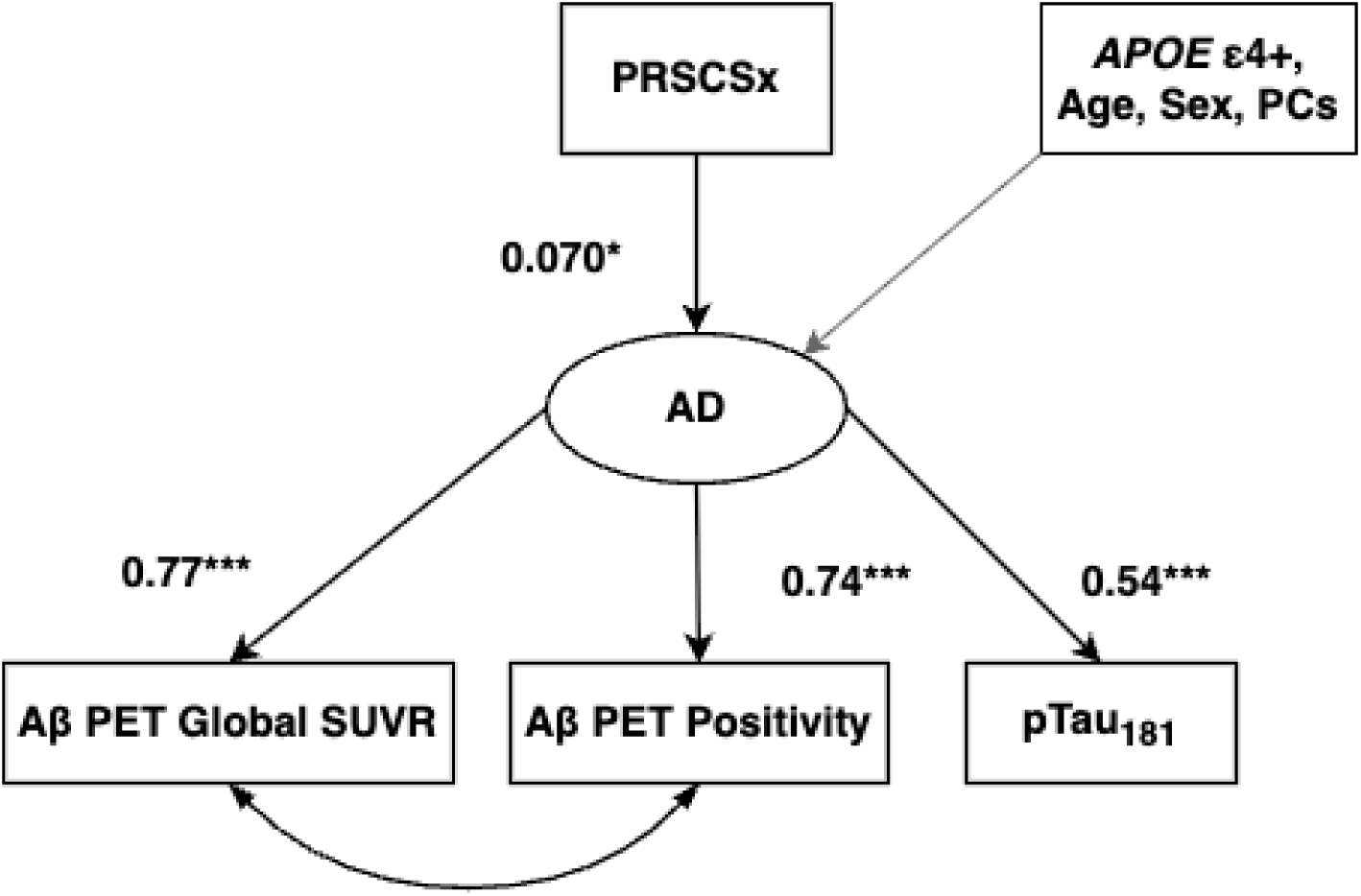
First-order path analysis of the association between PRS-CSx and the AD latent variable in the Health and Aging Brain Study - Health Disparities (HABSHD). N = 2,297, CFI = 0.983, TLI = 0.963, RMSEA = 0.037, SRMR = 0.021.

## DISCUSSION

In this study, we sought to evaluate the predictive accuracy of single-, multi-, and cross-ancestry AD-PRS models across individuals of African, Amerindian, and European ancestries to assess PRS transportability in multi-ancestral populations. We derived PRS models using multi-ancestry GWAS summary statistics and evaluated their predictive accuracy within the ADSP cohort. Our findings showed that the single-ancestry PRS showed the strongest predictive accuracy and AD risk associations in participants of European ancestry. However, its performance declined in non-European populations, particularly in participants of African ancestry, consistent with previous reports highlighting ancestry-specific limitations in PRS-based risk stratification^51–54^. This highlights reduced accuracy and portability of single-ancestry PRS model across ancestries, likely driven by differences in LD structure, allele frequency, and genetic architecture^55–59^. In contrast, multi- and cross-ancestry PRS models demonstrated improved predictive accuracy in African and Amerindian ancestry groups. When models incorporated a large number of SNPs from multi-ancestry GWAS (e.g., MAMA PT_0.1_, PRS-CSx), both associations with AD risk and explained variance (R²) improved, indicating that including more ancestry-matched SNPs capture better risk profiles in African and Admixed populations. In contrast, the absence of this trend for single-ancestry EUR PRS likely reflects the limited portability of European-derived variants to non-European populations with more variable genetic architectures. This supports previous findings that PRS constructed from smaller, ancestry-matched GWAS can outperform those derived from larger European datasets^25^. Although the non-European GWAS datasets used in this study were substantially smaller than the European GWAS dataset, incorporating base datasets that match the ancestry of the target populations improved overall PRS performance. In contrast, one study reported similar performance across single-, multi-, and cross-ancestry AD-PRS models using the MVP data^60^. This discrepancy may be due to differences in SNP selections. For example, their single-ancestry PRS included all GWS SNPs, whereas our approach applied LD pruning to select independent SNPs. As a result, incorporating additional SNPs from multi-ancestry GWAS in their framework may have contributed only marginal effects. Differences in cohort characteristics may also play a role as AD cases and controls in MVP were defined using ICD codes rather than clinical or biomarker-confirmed diagnoses.

Beyond AD diagnosis, we evaluated the clinical validity of PRS in predicting abnormalities in AD endophenotypes for Aβ, Tau, and cognitive function. The best-performing model, PRS-CSx, was associated with CSF Aβ_42_ levels and cognitive domains of memory, executive function, and language. Notably, PRS-CSx was not associated with CSF pTau_181_ or total tau, which may stem from sampling variability, data harmonization differences, and/or small sample sizes. Higher PRS-CSx was also associated with the most severe neuropathology burdens across CERAD, Thal phase, and Braak stage. In contrast, associations with lower or intermediate stages were weak or absent, suggesting that PRS-CSx predicts advanced pathology but may be less sensitive to earlier or moderate changes. In the HABS-HD cohort, PRS-CSx was significantly associated with an AD latent variable composed of plasma pTau_181_ and Aβ PET measures, while associations in single-outcome regression models did not remain significant after correction for multiple testing. This suggests that latent constructs may offer higher sensitivity and better capture biologically relevant effects. Across both regression and SEM analyses, limited sample sizes may have reduced power. In particular, the African ancestry group exhibited unstable parameter estimates, including a path loading exceeding 1, indicating potential model estimation instability due to small sample size.

Ancestry normalization emerged as a critical step in improving prediction accuracy for underrepresented groups by accounting for population-specific genetic structures that differ in allele frequency and LD. Although several PRS methods have been developed to include ancestry stratification or ancestry-specific SNP effects to address bias in cross-population prediction^22,23,30,61–63^, explicit normalization of the PRS itself based on global ancestry is still not widely implemented for AD-PRS. Shifts in PRS distributions that arise from differences in MAF and LD complicate the assessment of risks across populations and studies. By employing a reproducible framework such as the pgsc_calc Nextflow pipeline, genetic ancestry estimation and ancestry normalization can be performed effectively and seamlessly. Integrating ancestry normalization into PRS construction enables more equitable genetic prediction tools that better reflect the genetic architecture of ancestrally diverse populations. Given the high degree of genetic heterogeneity within a single superpopulation^64^ (e.g. African and admixed populations), ancestry normalization is a critical step to account for differential risk profiles that arise from ancestral differences rather than true genetic risk.

Several limitations should be acknowledged in our study. First, a lack of representation of people of South and East Asian ancestry limited our ability to assess the cross-ancestry transportability of the PRS models. The dataset is also limited to participants in the US, and how multi- and cross-ancestry perform in non-European populations from other continents is unknown. Furthermore, the currently available GWAS summary statistics from minority populations still don’t reflect the full genetic diversity and substructures that exist within one superpopulation. This calls for the necessities to continue expanding and improving the diversity and sample sizes of future GWAS to more fully represent global and intra-ancestry genetic variation. Second, we did not incorporate genotype–environment (GxE) interactions, a framework that explains the interplay of environmental, socioeconomic, and lifestyle factors that can modify genetic risk. These factors are known to contribute additively or multiplicatively to AD susceptibility across populations^65–67^. Third, future iterations of such studies should prioritize larger, more representative cohorts and integrate extensive phenotypic data from biobanks (e.g., All of Us, UK biobanks) to enhance both the calibration and clinical adaptability of risk predictions. Finally, the application of other advanced data-driven models, such as LDpred2^68^, PolyPred+^69^, and BridgPRS^22,70^, and incorporation of SNP heritability estimates could further refine the predictive power and cross-population transferability of AD-PRS^71^. With the rapid proliferation of PRS methods^72^, future studies may focus on benchmarking across all the methods in large, diverse cohorts as well as community-based populations.

Despite these limitations, our study demonstrates several key strengths. We systematically evaluated single-, multi-, and cross-ancestry PRS models across participants of African, Amerindian, and European ancestries, providing a comprehensive assessment of PRS transferability in diverse cohorts. By incorporating ancestry-specific normalization and leveraging multi-ancestry GWAS summary statistics, we improved predictive accuracy in underrepresented populations and highlighted the importance of including ancestry-matched SNPs in risk models. Furthermore, by extending the evaluation beyond AD diagnosis to biomarkers and cognitive endophenotypes within the A/T/N framework, we provide evidence for the clinical validity of PRS in capturing biologically relevant AD pathology. The broader implications of our findings align with the objectives of the eMERGE Consortium’s Genome Informed Risk Assessment (GIRA) initiative, which emphasizes the integration of PRS into clinical care while ensuring analytical validity and health equity across diverse populations^73^. By leveraging diverse datasets and applying ancestry-specific normalization to PRS calculations, our methodology enhances risk stratification accuracy, supporting the growing consensus that such approaches are crucial for equitable and effective genomic medicine^16,20,21,23,62,74–76^.

In summary, this study highlights methodological innovations of leveraging multi-ancestry base GWAS summary statistics, applying ancestry normalization, and extending evaluation beyond diagnosis to AD endophenotypes that can enhance both the accuracy and clinical relevance of AD-PRS. Importantly, our results call for equitable genomic medicine through increasing genetic representation and moving beyond Eurocentric risk models to reflect the full spectrum of human genetic diversity. By demonstrating practical strategies to improve prediction in underrepresented populations, this work provides a foundation for more inclusive risk assessment tools that can be translated into meaningful clinical applications.

## CONSENT STATEMENT

## Acknowledgments

The authors would like to thank all the participants, staff, researcher teams, and partners of the Alzheimer’s Disease Sequencing Project (ADSP). The ADSP is comprised of two Alzheimer’s Disease (AD) genetics consortia and three National Human Genome Research Institute (NHGRI) funded Large Scale Sequencing and Analysis Centers (LSAC). The two AD genetics consortia are the Alzheimer’s Disease Genetics Consortium (ADGC) funded by NIA (U01 AG032984), and the Cohorts for Heart and Aging Research in Genomic Epidemiology (CHARGE) funded by NIA (R01 AG033193), the National Heart, Lung, and Blood Institute (NHLBI), other National Institute of Health (NIH) institutes and other foreign governmental and non-governmental organizations. The Discovery Phase analysis of sequence data is supported through UF1AG047133 (to Drs. Schellenberg, Farrer, Pericak-Vance, Mayeux, and Haines); U01AG049505 to Dr. Seshadri; U01AG049506 to Dr. Boerwinkle; U01AG049507 to Dr. Wijsman; and U01AG049508 to Dr. Goate and the Discovery Extension Phase analysis is supported through U01AG052411 to Dr. Goate, U01AG052410 to Dr. Pericak-Vance and U01 AG052409 to Drs. Seshadri and Fornage.

Sequencing for the Follow Up Study (FUS) is supported through U01AG057659 (to Drs. PericakVance, Mayeux, and Vardarajan) and U01AG062943 (to Drs. Pericak-Vance and Mayeux). Data generation and harmonization in the Follow-up Phase is supported by U54AG052427 (to Drs. Schellenberg and Wang). The FUS Phase analysis of sequence data is supported through U01AG058589 (to Drs. Destefano, Boerwinkle, De Jager, Fornage, Seshadri, and Wijsman), U01AG058654 (to Drs. Haines, Bush, Farrer, Martin, and Pericak-Vance), U01AG058635 (to Dr. Goate), RF1AG058066 (to Drs. Haines, Pericak-Vance, and Scott), RF1AG057519 (to Drs. Farrer and Jun), R01AG048927 (to Dr. Farrer), and RF1AG054074 (to Drs. Pericak-Vance and Beecham).

The ADGC cohorts include: Adult Changes in Thought (ACT) (U01 AG006781, U19 AG066567), the Alzheimer’s Disease Research Centers (ADRC) (P30 AG062429, P30 AG066468, P30 AG062421, P30 AG066509, P30 AG066514, P30 AG066530, P30 AG066507, P30 AG066444, P30 AG066518, P30 AG066512, P30 AG066462, P30 AG072979, P30 AG072972, P30 AG072976, P30 AG072975, P30 AG072978, P30 AG072977, P30 AG066519, P30 AG062677, P30 AG079280, P30 AG062422, P30 AG066511, P30 AG072946, P30 AG062715, P30 AG072973, P30 AG066506, P30 AG066508, P30 AG066515, P30 AG072947, P30 AG072931, P30 AG066546, P20 AG068024, P20 AG068053, P20 AG068077, P20 AG068082, P30 AG072958, P30 AG072959), the Chicago Health and Aging Project (CHAP) (R01 AG11101, RC4 AG039085, K23 AG030944), Indiana Memory and Aging Study (IMAS) (R01 AG019771), Indianapolis Ibadan (R01 AG009956, P30 AG010133), the Memory and Aging Project (MAP) (R01 AG17917), Mayo Clinic (MAYO) (R01 AG032990, U01 AG046139, R01 NS080820, RF1 AG051504, P50 AG016574), Mayo Parkinson’s Disease controls (NS039764, NS071674, 5RC2HG005605), University of Miami (R01 AG027944, R01 AG028786, R01 AG019085, IIRG09133827, A2011048), the Multi-Institutional Research in Alzheimer’s Genetic Epidemiology Study (MIRAGE) (R01 AG09029, R01 AG025259), the National Centralized Repository for Alzheimer’s Disease and Related Dementias (NCRAD) (U24 AG021886), the National Institute on Aging Late Onset Alzheimer’s Disease Family Study (NIA-LOAD) (U24 AG056270), the Religious Orders Study (ROS) (P30 AG10161, R01 AG15819), the Texas Alzheimer’s Research and Care Consortium (TARCC) (funded by the Darrell K Royal Texas Alzheimer’s Initiative), Vanderbilt University/Case Western Reserve University (VAN/CWRU) (R01 AG019757, R01 AG021547, R01 AG027944, R01 AG028786, P01 NS026630, and Alzheimer’s Association), the Washington Heights-Inwood Columbia Aging Project (WHICAP) (RF1 AG054023), the University of Washington Families (VA Research Merit Grant, NIA: P50AG005136, R01AG041797, NINDS: R01NS069719), the Columbia University Hispanic Estudio Familiar de Influencia Genetica de Alzheimer (EFIGA) (RF1 AG015473), the University of Toronto (UT) (funded by Wellcome Trust, Medical Research Council, Canadian Institutes of Health Research), and Genetic Differences (GD) (R01 AG007584). The CHARGE cohorts are supported in part by National Heart, Lung, and Blood Institute (NHLBI) infrastructure grant HL105756 (Psaty), RC2HL102419 (Boerwinkle) and the neurology working group is supported by the National Institute on Aging (NIA) R01 grant AG033193.

The CHARGE cohorts participating in the ADSP include the following: Austrian Stroke Prevention Study (ASPS), ASPS-Family study, and the Prospective Dementia Registry-Austria (ASPS/PRODEM-Aus), the Atherosclerosis Risk in Communities (ARIC) Study, the Cardiovascular Health Study (CHS), the Erasmus Rucphen Family Study (ERF), the Framingham Heart Study (FHS), and the Rotterdam Study (RS). ASPS is funded by the Austrian Science Fond (FWF) grant number P20545-P05 and P13180 and the Medical University of Graz. The ASPS-Fam is funded by the Austrian Science Fund (FWF) project I904), the EU Joint Programme – Neurodegenerative Disease Research (JPND) in frame of the BRIDGET project (Austria, Ministry of Science) and the Medical University of Graz and the Steiermärkische Krankenanstalten Gesellschaft. PRODEM-Austria is supported by the Austrian Research Promotion agency (FFG) (Project No. 827462) and by the Austrian National Bank (Anniversary Fund, project 15435. ARIC research is carried out as a collaborative study supported by NHLBI contracts (HHSN268201100005C, HHSN268201100006C, HHSN268201100007C, HHSN268201100008C, HHSN268201100009C, HHSN268201100010C, HHSN268201100011C, and HHSN268201100012C). Neurocognitive data in ARIC is collected by U01 2U01HL096812, 2U01HL096814, 2U01HL096899, 2U01HL096902, 2U01HL096917 from the NIH (NHLBI, NINDS, NIA and NIDCD), and with previous brain MRI examinations funded by R01-HL70825 from the NHLBI. CHS research was supported by contracts HHSN268201200036C, HHSN268200800007C, N01HC55222, N01HC85079, N01HC85080, N01HC85081, N01HC85082, N01HC85083, N01HC85086, and grants U01HL080295 and U01HL130114 from the NHLBI with additional contribution from the National Institute of Neurological Disorders and Stroke (NINDS). Additional support was provided by R01AG023629, R01AG15928, and R01AG20098 from the NIA. FHS research is supported by NHLBI contracts N01-HC-25195 and HHSN268201500001I. This study was also supported by additional grants from the NIA (R01s AG054076, AG049607 and AG033040 and NINDS (R01 NS017950). The ERF study as a part of EUROSPAN (European Special Populations Research Network) was supported by European Commission FP6 STRP grant number 018947 (LSHG-CT-2006-01947) and also received funding from the European Community’s Seventh Framework Programme (FP7/2007-2013)/grant agreement HEALTH-F4-2007-201413 by the European Commission under the programme “Quality of Life and Management of the Living Resources” of 5th Framework Programme (no. QLG2-CT-2002-01254). High-throughput analysis of the ERF data was supported by a joint grant from the Netherlands Organization for Scientific Research and the Russian Foundation for Basic Research (NWO-RFBR 047.017.043). The Rotterdam Study is funded by Erasmus Medical Center and Erasmus University, Rotterdam, the Netherlands Organization for Health Research and Development (ZonMw), the Research Institute for Diseases in the Elderly (RIDE), the Ministry of Education, Culture and Science, the Ministry for Health, Welfare and Sports, the European Commission (DG XII), and the municipality of Rotterdam. Genetic data sets are also supported by the Netherlands Organization of Scientific Research NWO Investments (175.010.2005.011, 911-03-012), the Genetic Laboratory of the Department of Internal Medicine, Erasmus MC, the Research Institute for Diseases in the Elderly (014-93-015; RIDE2), and the Netherlands Genomics Initiative (NGI)/Netherlands Organization for Scientific Research (NWO) Netherlands Consortium for Healthy Aging (NCHA), project 050-060-810. All studies are grateful to their participants, faculty and staff. The content of these manuscripts is solely the responsibility of the authors and does not necessarily represent the official views of the National Institutes of Health or the U.S. Department of Health and Human Services.

The FUS cohorts include: the Alzheimer’s Disease Research Centers (ADRC) (P30 AG062429, P30 AG066468, P30 AG062421, P30 AG066509, P30 AG066514, P30 AG066530, P30 AG066507, P30 AG066444, P30 AG066518, P30 AG066512, P30 AG066462, P30 AG072979, P30 AG072972, P30 AG072976, P30 AG072975, P30 AG072978, P30 AG072977, P30 AG066519, P30 AG062677, P30 AG079280, P30 AG062422, P30 AG066511, P30 AG072946, P30 AG062715, P30 AG072973, P30 AG066506, P30 AG066508, P30 AG066515, P30 AG072947, P30 AG072931, P30 AG066546, P20 AG068024, P20 AG068053, P20 AG068077, P20 AG068082, P30 AG072958, P30 AG072959), Alzheimer’s Disease Neuroimaging Initiative (ADNI) (U19AG024904), Amish Protective Variant Study (RF1AG058066), Cache County Study (R01AG11380, R01AG031272, R01AG21136, RF1AG054052), Case Western Reserve University Brain Bank (CWRUBB) (P50AG008012), Case Western Reserve University Rapid Decline (CWRURD) (RF1AG058267, NU38CK000480), CubanAmerican Alzheimer’s Disease Initiative (CuAADI) (3U01AG052410), Estudio Familiar de Influencia Genetica en Alzheimer (EFIGA) (5R37AG015473, RF1AG015473, R56AG051876), Genetic and Environmental Risk Factors for Alzheimer Disease Among African Americans Study (GenerAAtions) (2R01AG09029, R01AG025259, 2R01AG048927), Gwangju Alzheimer and Related Dementias Study (GARD) (U01AG062602), Hillblom Aging Network (2014-A-004-NET, R01AG032289, R01AG048234), Hussman Institute for Human Genomics Brain Bank (HIHGBB) (R01AG027944, Alzheimer’s Association “Identification of Rare Variants in Alzheimer Disease”), Ibadan Study of Aging (IBADAN) (5R01AG009956), Longevity Genes Project (LGP) and LonGenity (R01AG042188, R01AG044829, R01AG046949, R01AG057909, R01AG061155, P30AG038072), Mexican Health and Aging Study (MHAS) (R01AG018016), Multi-Institutional Research in Alzheimer’s Genetic Epidemiology (MIRAGE) (2R01AG09029, R01AG025259, 2R01AG048927), Northern Manhattan Study (NOMAS) (R01NS29993), Peru Alzheimer’s Disease Initiative (PeADI) (RF1AG054074), Puerto Rican 1066 (PR1066) (Wellcome Trust (GR066133/GR080002), European Research Council (340755)), Puerto Rican Alzheimer Disease Initiative (PRADI) (RF1AG054074), Reasons for Geographic and Racial Differences in Stroke (REGARDS) (U01NS041588), Research in African American Alzheimer Disease Initiative (REAAADI) (U01AG052410), the Religious Orders Study (ROS) (P30 AG10161, P30 AG72975, R01 AG15819, R01 AG42210), the RUSH Memory and Aging Project (MAP) (R01 AG017917, R01 AG42210Stanford Extreme Phenotypes in AD (R01AG060747), University of Miami Brain Endowment Bank (MBB), University of Miami/Case Western/North Carolina A&T African American (UM/CASE/NCAT) (U01AG052410, R01AG028786), and Wisconsin Registry for Alzheimer’s Prevention (WRAP) (R01AG027161 and R01AG054047).

The four LSACs are: the Human Genome Sequencing Center at the Baylor College of Medicine (U54 HG003273), the Broad Institute Genome Center (U54HG003067), The American Genome Center at the Uniformed Services University of the Health Sciences (U01AG057659), and the Washington University Genome Institute (U54HG003079). Genotyping and sequencing for the ADSP FUS is also conducted at John P. Hussman Institute for Human Genomics (HIHG) Center for Genome Technology (CGT).

Biological samples and associated phenotypic data used in primary data analyses were stored at Study Investigators institutions, and at the National Centralized Repository for Alzheimer’s Disease and Related Dementias (NCRAD, U24AG021886) at Indiana University funded by NIA. Associated Phenotypic Data used in primary and secondary data analyses were provided by Study Investigators, the NIA funded Alzheimer’s Disease Centers (ADCs), and the National Alzheimer’s Coordinating Center (NACC, U24AG072122) and the National Institute on Aging Genetics of Alzheimer’s Disease Data Storage Site (NIAGADS, U24AG041689) at the University of Pennsylvania, funded by NIA. Harmonized phenotypes were provided by the ADSP Phenotype Harmonization Consortium (ADSP-PHC), funded by NIA (U24 AG074855, U01 AG068057 and R01 AG059716) and Ultrascale Machine Learning to Empower Discovery in Alzheimer’s Disease Biobanks (AI4AD, U01 AG068057). This research was supported in part by the Intramural Research Program of the National Institutes of health, National Library of Medicine. Contributors to the Genetic Analysis Data included Study Investigators on projects that were individually funded by NIA, and other NIH institutes, and by private U.S. organizations, or foreign governmental or nongovernmental organizations.

The ADSP Phenotype Harmonization Consortium (ADSP-PHC) is funded by NIA (U24 AG074855, U01 AG068057 and R01 AG059716). The harmonized cohorts within the ADSP-PHC include: the Anti-Amyloid Treatment in Asymptomatic Alzheimer’s study (A4 Study), a secondary prevention trial in preclinical Alzheimer’s disease, aiming to slow cognitive decline associated with brain amyloid accumulation in clinically normal older individuals. The A4 Study is funded by a public-private-philanthropic partnership, including funding from the National Institutes of Health-National Institute on Aging, Eli Lilly and Company, Alzheimer’s Association, Accelerating Medicines Partnership, GHR Foundation, an anonymous foundation and additional private donors, with in-kind support from Avid and Cogstate. The companion observational Longitudinal Evaluation of Amyloid Risk and Neurodegeneration (LEARN) Study is funded by the Alzheimer’s Association and GHR Foundation. The A4 and LEARN Studies are led by Dr. Reisa Sperling at Brigham and Women’s Hospital, Harvard Medical School and Dr. Paul Aisen at the Alzheimer’s Therapeutic Research Institute (ATRI), University of Southern California. The A4 and LEARN Studies are coordinated by ATRI at the University of Southern California, and the data are made available through the Laboratory for Neuro Imaging at the University of Southern California. The participants screening for the A4 Study provided permission to share their de-identified data in order to advance the quest to find a successful treatment for Alzheimer’s disease. We would like to acknowledge the dedication of all the participants, the site personnel, and all of the partnership team members who continue to make the A4 and LEARN Studies possible. The complete A4 Study Team list is available on: a4study.org/a4-study-team.; the Adult Changes in Thought study (ACT), U01 AG006781, U19 AG066567; Alzheimer’s Disease Neuroimaging Initiative (ADNI): Data collection and sharing for this project was funded by the Alzheimer’s Disease Neuroimaging Initiative (ADNI) (National Institutes of Health Grant U01 AG024904) and DOD ADNI (Department of Defense award number W81XWH-12-2-0012). ADNI is funded by the National Institute on Aging, the National Institute of Biomedical Imaging and Bioengineering, and through generous contributions from the following: AbbVie, Alzheimer’s Association; Alzheimer’s Drug Discovery Foundation; Araclon Biotech; BioClinica, Inc.; Biogen; Bristol-Myers Squibb Company; CereSpir, Inc.; Cogstate; Eisai Inc.; Elan Pharmaceuticals, Inc.; Eli Lilly and Company; EuroImmun; F. Hoffmann-La Roche Ltd and its affiliated company Genentech, Inc.; Fujirebio; GE Healthcare; IXICO Ltd.;Janssen Alzheimer Immunotherapy Research & Development, LLC.; Johnson & Johnson Pharmaceutical Research & Development LLC.; Lumosity; Lundbeck; Merck & Co., Inc.;Meso Scale Diagnostics, LLC.; NeuroRx Research; Neurotrack Technologies; Novartis Pharmaceuticals Corporation; Pfizer Inc.; Piramal Imaging; Servier; Takeda Pharmaceutical Company; and Transition Therapeutics. The Canadian Institutes of Health Research is providing funds to support ADNI clinical sites in Canada. Private sector contributions are facilitated by the Foundation for the National Institutes of Health (www.fnih.org). The grantee organization is the Northern California Institute for Research and Education, and the study is coordinated by the Alzheimer’s Therapeutic Research Institute at the University of Southern California. ADNI data are disseminated by the Laboratory for Neuro Imaging at the University of Southern California; Estudio Familiar de Influencia Genetica en Alzheimer (EFIGA): 5R37AG015473, RF1AG015473, R56AG051876; Memory & Aging Project at Knight Alzheimer’s Disease Research Center (MAP at Knight ADRC): The Memory and Aging Project at the Knight-ADRC (Knight-ADRC). This work was supported by the National Institutes of Health (NIH) grants R01AG064614, R01AG044546, RF1AG053303, RF1AG058501, U01AG058922 and R01AG064877 to Carlos Cruchaga. The recruitment and clinical characterization of research participants at Washington University was supported by NIH grants P30AG066444, P01AG03991, and P01AG026276. Data collection and sharing for this project was supported by NIH grants RF1AG054080, P30AG066462, R01AG064614 and U01AG052410. We thank the contributors who collected samples used in this study, as well as patients and their families, whose help and participation made this work possible. This work was supported by access to equipment made possible by the Hope Center for Neurological Disorders, the Neurogenomics and Informatics Center (NGI: https://neurogenomics.wustl.edu/) and the Departments of Neurology and Psychiatry at Washington University School of Medicine; National Alzheimer’s Coordinating Center (NACC): The NACC database is funded by NIA/NIH Grant U24 AG072122. NACC data are contributed by the NIA-funded ADRCs: P30 AG062429 (PI James Brewer, MD, PhD), P30 AG066468 (PI Oscar Lopez, MD), P30 AG062421 (PI Bradley Hyman, MD, PhD), P30 AG066509 (PI Thomas Grabowski, MD), P30 AG066514 (PI Mary Sano, PhD), P30 AG066530 (PI Helena Chui, MD), P30 AG066507 (PI Marilyn Albert, PhD), P30 AG066444 (PI John Morris, MD), P30 AG066518 (PI Jeffrey Kaye, MD), P30 AG066512 (PI Thomas Wisniewski, MD), P30 AG066462 (PI Scott Small, MD), P30 AG072979 (PI David Wolk, MD), P30 AG072972 (PI Charles DeCarli, MD), P30 AG072976 (PI Andrew Saykin, PsyD), P30 AG072975 (PI David Bennett, MD), P30 AG072978 (PI Neil Kowall, MD), P30 AG072977 (PI Robert Vassar, PhD), P30 AG066519 (PI Frank LaFerla, PhD), P30 AG062677 (PI Ronald Petersen, MD, PhD), P30 AG079280 (PI Eric Reiman, MD), P30 AG062422 (PI Gil Rabinovici, MD), P30 AG066511 (PI Allan Levey, MD, PhD), P30 AG072946 (PI Linda Van Eldik, PhD), P30 AG062715 (PI Sanjay Asthana, MD, FRCP), P30 AG072973 (PI Russell Swerdlow, MD), P30 AG066506 (PI Todd Golde, MD, PhD), P30 AG066508 (PI Stephen Strittmatter, MD, PhD), P30 AG066515 (PI Victor Henderson, MD, MS), P30 AG072947 (PI Suzanne Craft, PhD), P30 AG072931 (PI Henry Paulson, MD, PhD), P30 AG066546 (PI Sudha Seshadri, MD), P20 AG068024 (PI Erik Roberson, MD, PhD), P20 AG068053 (PI Justin Miller, PhD), P20 AG068077 (PI Gary Rosenberg, MD), P20 AG068082 (PI Angela Jefferson, PhD), P30 AG072958 (PI Heather Whitson, MD), P30 AG072959 (PI James Leverenz, MD); National Institute on Aging Alzheimer’s Disease Family Based Study (NIA-AD FBS): U24 AG056270; Religious Orders Study (ROS): P30AG10161,R01AG15819, R01AG42210; Memory and Aging Project (MAP - Rush): R01AG017917, R01AG42210; Minority Aging Research Study (MARS): R01AG22018, R01AG42210; Washington Heights/Inwood Columbia Aging Project (WHICAP): RF1 AG054023;and Wisconsin Registry for Alzheimer’s Prevention (WRAP): R01AG027161 and R01AG054047. Additional acknowledgments include the National Institute on Aging Genetics of Alzheimer’s Disease Data Storage Site (NIAGADS, U24AG041689) at the University of Pennsylvania, funded by NIA.

Data for this study were prepared, archived, and distributed by the National Institute on Aging Alzheimer’s Disease Data Storage Site (NIAGADS) at the University of Pennsylvania (U24-AG041689), funded by the National Institute on Aging.

The NACC database is funded by NIA/NIH Grant U24 AG072122. NACC data are contributed by the NIA-funded ADRCs: P30 AG062429 (PI James Brewer, MD, PhD), P30 AG066468 (PI Oscar Lopez, MD), P30 AG062421 (PI Bradley Hyman, MD, PhD), P30 AG066509 (PI Thomas Grabowski, MD), P30 AG066514 (PI Mary Sano, PhD), P30 AG066530 (PI Helena Chui, MD), P30 AG066507 (PI Marilyn Albert, PhD), P30 AG066444 (PI John Morris, MD), P30 AG066518 (PI Jeffrey Kaye, MD), P30 AG066512 (PI Thomas Wisniewski, MD), P30 AG066462 (PI Scott Small, MD), P30 AG072979 (PI David Wolk, MD), P30 AG072972 (PI Charles DeCarli, MD), P30 AG072976 (PI Andrew Saykin, PsyD), P30 AG072975 (PI David Bennett, MD), P30 AG072978 (PI Neil Kowall, MD), P30 AG072977 (PI Robert Vassar, PhD), P30 AG066519 (PI Frank LaFerla, PhD), P30 AG062677 (PI Ronald Petersen, MD, PhD), P30 AG079280 (PI Eric Reiman, MD), P30 AG062422 (PI Gil Rabinovici, MD), P30 AG066511 (PI Allan Levey, MD, PhD), P30 AG072946 (PI Linda Van Eldik, PhD), P30 AG062715 (PI Sanjay Asthana, MD, FRCP), P30 AG072973 (PI Russell Swerdlow, MD), P30 AG066506 (PI Todd Golde, MD, PhD), P30 AG066508 (PI Stephen Strittmatter, MD, PhD), P30 AG066515 (PI Victor Henderson, MD, MS), P30 AG072947 (PI Suzanne Craft, PhD), P30 AG072931 (PI Henry Paulson, MD, PhD), P30 AG066546 (PI Sudha Seshadri, MD), P20 AG068024 (PI Erik Roberson, MD, PhD), P20 AG068053 (PI Justin Miller, PhD), P20 AG068077 (PI Gary Rosenberg, MD), P20 AG068082 (PI Angela Jefferson, PhD), P30 AG072958 (PI Heather Whitson, MD), P30 AG072959 (PI James Leverenz, MD).

Data collection and sharing for this project was funded by the Alzheimer’s Disease Neuroimaging Initiative (ADNI) (National Institutes of Health Grant U01 AG024904) and DOD ADNI (Department of Defense award number W81XWH-12-2-0012). ADNI is funded by the National Institute on Aging, the National Institute of Biomedical Imaging and Bioengineering, and through generous contributions from the following: AbbVie, Alzheimer’s Association; Alzheimer’s Drug Discovery Foundation; Araclon Biotech; BioClinica, Inc.; Biogen; Bristol-Myers Squibb Company; CereSpir, Inc.; Cogstate; Eisai Inc.; Elan Pharmaceuticals, Inc.; Eli Lilly and Company; EuroImmun; F. Hoffmann-La Roche Ltd and its affiliated company Genentech, Inc.; Fujirebio; GE Healthcare; IXICO Ltd.;Janssen Alzheimer Immunotherapy Research & Development, LLC.; Johnson & Johnson Pharmaceutical Research & Development LLC.; Lumosity; Lundbeck; Merck & Co., Inc.;Meso Scale Diagnostics, LLC.; NeuroRx Research; Neurotrack Technologies; Novartis Pharmaceuticals Corporation; Pfizer Inc.; Piramal Imaging; Servier; Takeda Pharmaceutical Company; and Transition Therapeutics. The Canadian Institutes of Health Research is providing funds to support ADNI clinical sites in Canada. Private sector contributions are facilitated by the Foundation for the National Institutes of Health (www.fnih.org). The grantee organization is the Northern California Institute for Research and Education, and the study is coordinated by the Alzheimer’s Therapeutic Research Institute at the University of Southern California. ADNI data are disseminated by the Laboratory for Neuro Imaging at the University of Southern California.

The authors would like to thank all the participants, staff, researcher teams, and partners of the Health & Aging Brain Study - Health Disparities (HABS-HD). The HABS-HD is supported by the National Institute on Aging of the National Institutes of Health under Award Numbers R01AG054073, R01AG058533, R01AG070862, P41EB015922, and U19AG078109. The content is solely the responsibility of the authors and does not necessarily represent the official views of the National Institutes of Health. We gratefully acknowledge the contributions of our study partners and their families, whose help and participation made this work possible.

## DATA AVAILABILITY STATEMENT

European summary statistics from Bellenguez et al. 2022^5^ were accessed through the National Human Genome Research Institute-European Bioinformatics Institute using accession number GCST90027158. Finnish European summary statistics from FinnGen Release 6 were accessed through https://www.finngen.fi/en/access_results. African summary Statistics from Kunkle et al. 2021^28^ were accessed through NIAGADS (https://www.niagads.org/) using accession number NG00100. East Asian summary statistics from Shigemizu et al. 2021^29^ were accessed through the National Bioscience Database Center (NBDC) at the Japan Science and Technology Agency (JST) at https://humandbs.biosciencedbc.jp/en/ through accession number hum0237.v1.gwas.v1. Caribbean Hispanics summary statistics from the Columbia University Study of Caribbean Hispanics and Late-Onset Alzheimer’s disease were accessed via application to dbGaP accession number phs000496.v1.p1.

This work was conducted using the National Alzheimer’s Coordinating Center Uniform Dataset under application 10238; the Alzheimer’s Disease Neuroimaging Initiative under application SJA; and the Alzheimer’s Disease Sequencing Project under application 10050.

## CODE AVAILABILITY

All codes developed and used are available at https://github.com/AndrewsLabUCSF/AFRPRS/tree/main. Information regarding all the software and reference datasets used in the analysis can also be found in the repository.

