## Supplementary Figures 1-6 for "Cross-Ancestry Polygenic Risk Scores Enhance Alzheimer’s Disease Risk Prediction in Multiethnic Cohorts"

### SUPPLEMENTAL FIGURES

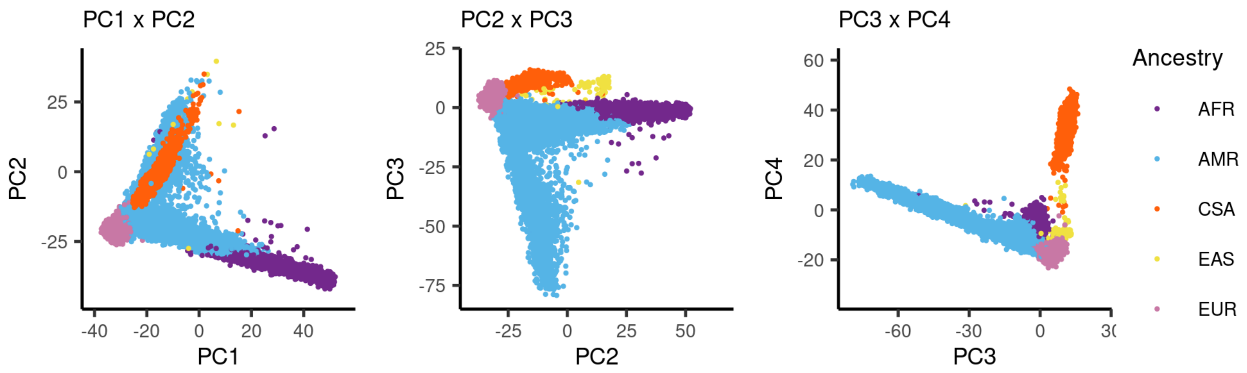


Supplementary Figure 1. Principal component analysis (PCA) of genetic data reveals global population structure. Scatterplots show the first four principal components (PCs) of genotype data: (left) PC1 vs PC2, (center) PC2 vs PC3, and (right) PC3 vs PC4. Each point represents an individual, colored by population group or inferred ancestry. Clustering patterns reflect genetic differentiation among major continental populations.

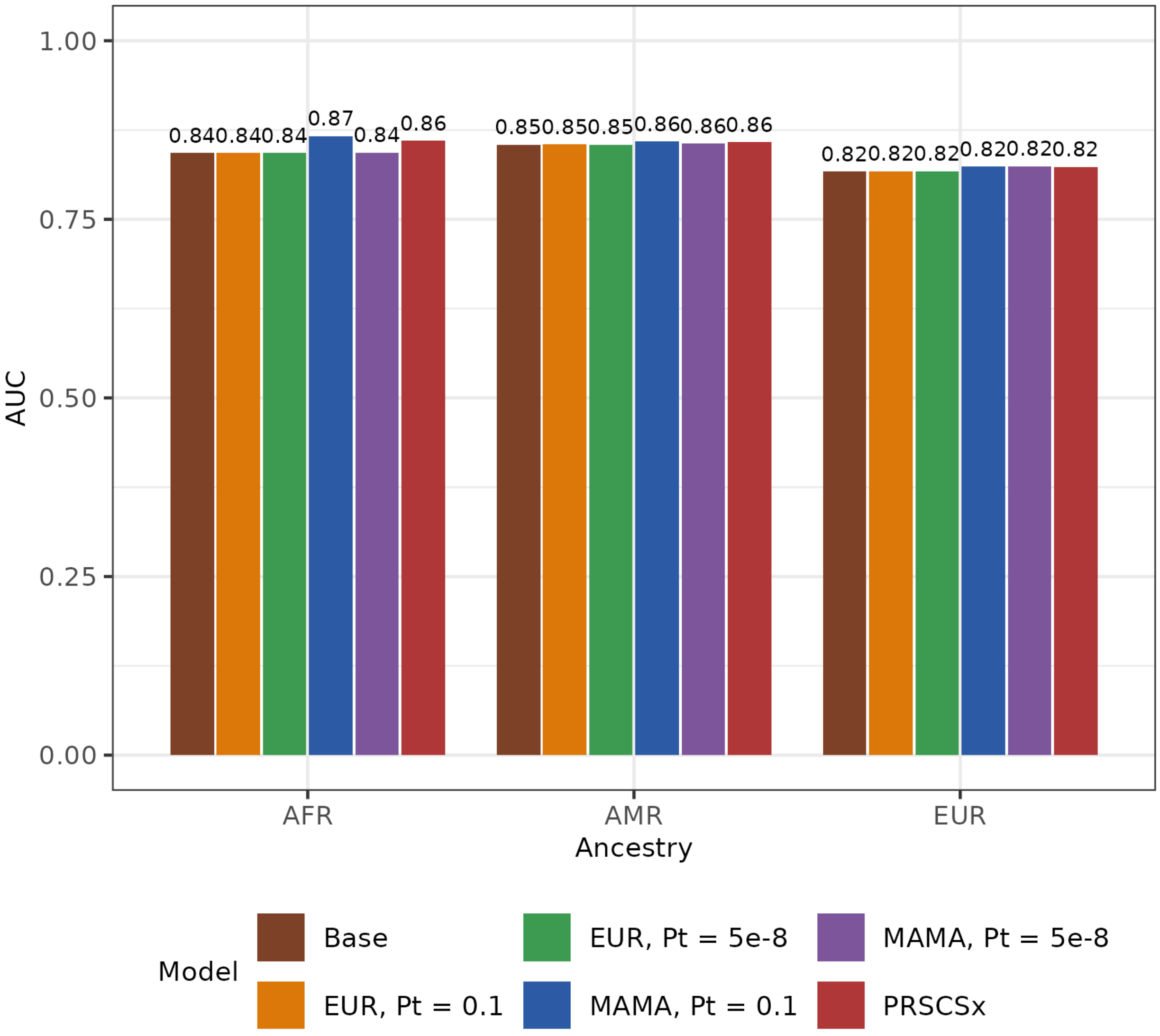


Supplementary Figure 2: AUC of ancestry-normalized single- (EUR), multi- (MAMA), and cross-ancestry (PRS-CSx) PRS models.

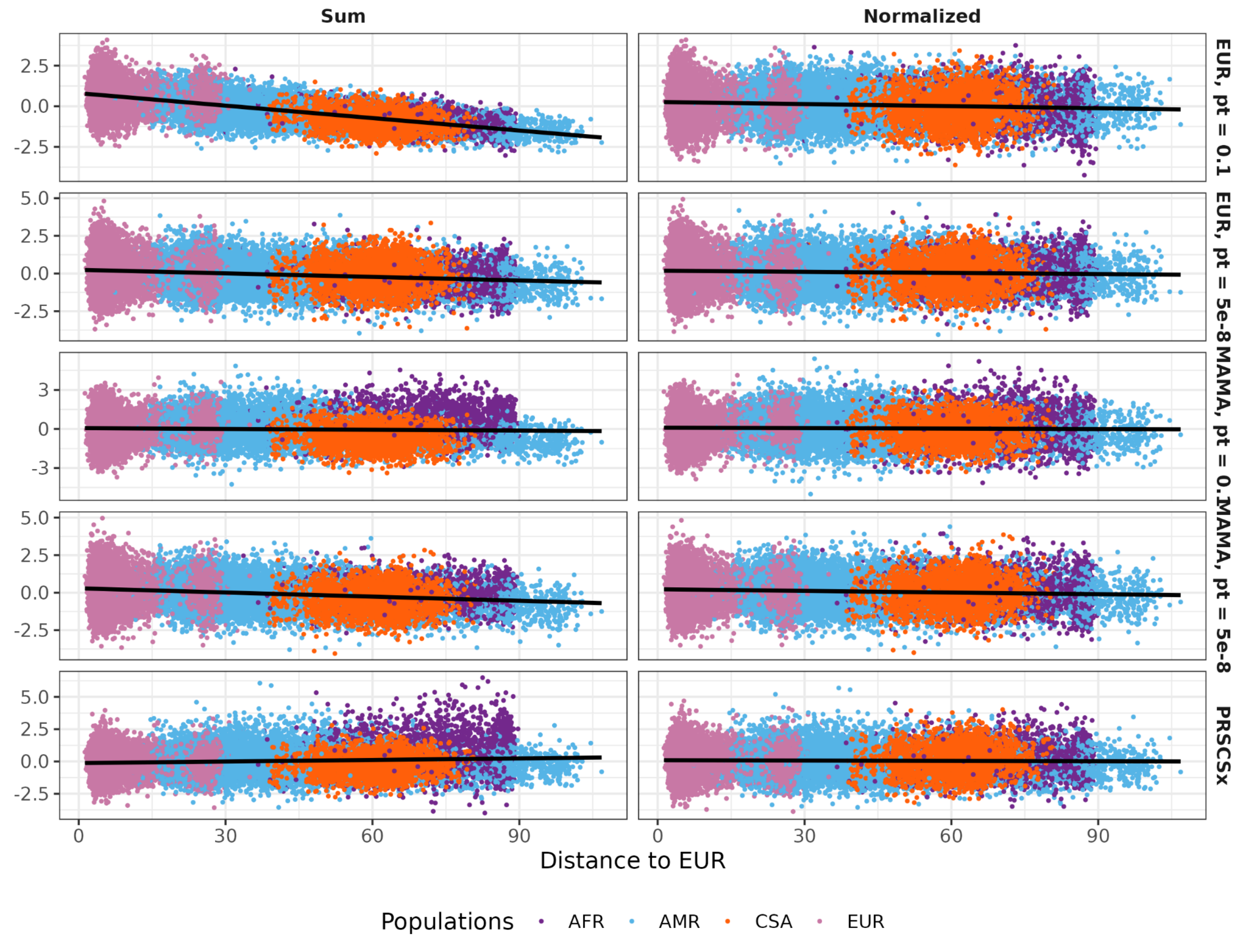


Supplementary Figure 3: Genetic distance analysis of ancestry for the ADSP cohort, with PRS score on the y-axis. “Sum” represents PRS prior to ancestry normalization, and “Normalized” represents PRS after the ancestry normalization.

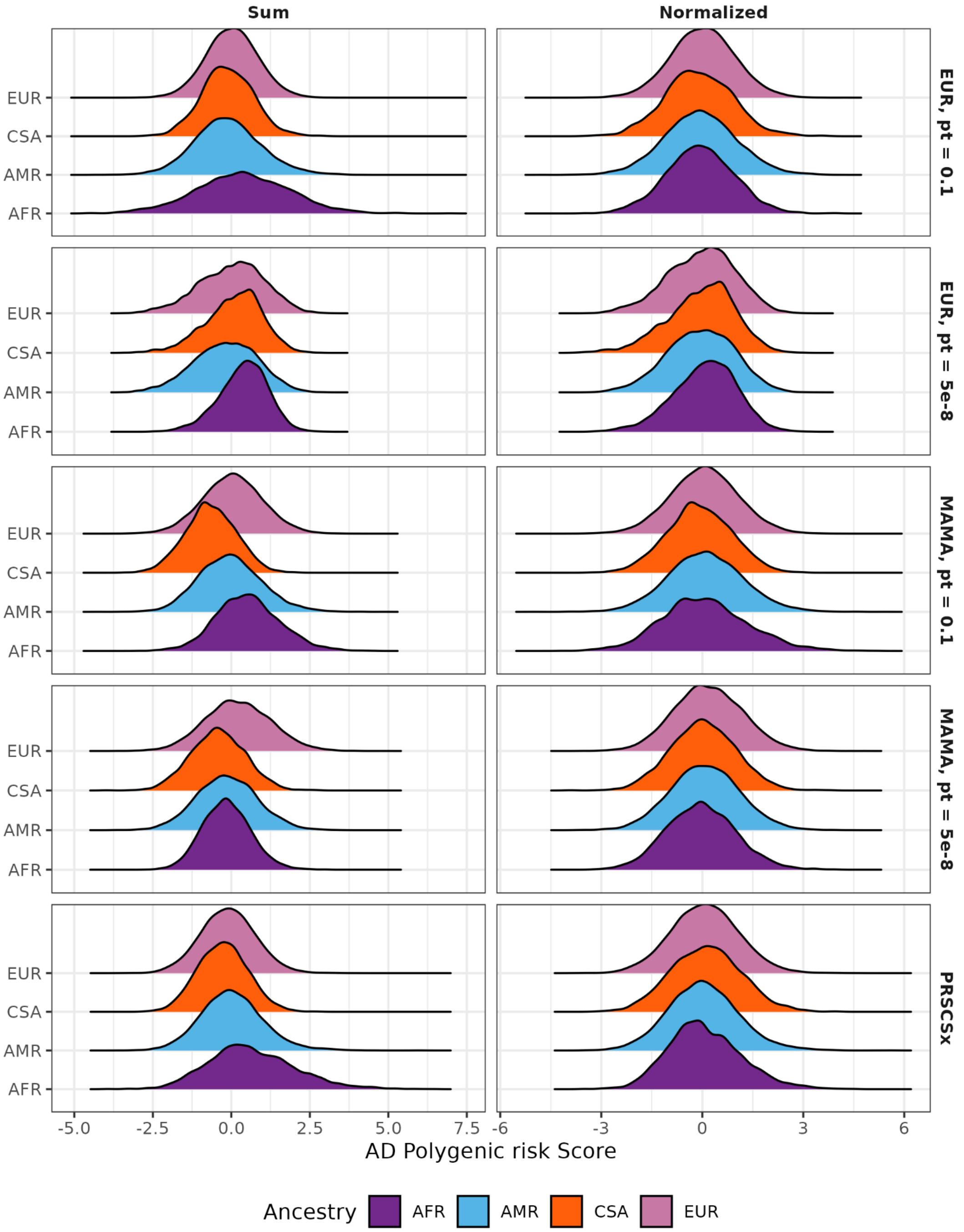


Supplementary Figure 4: Ancestry normalization of raw PRS performed by pgsc_calc. “sum” indicates raw PRS prior to ancestry normalization, and “z_norm2” indicates normalized PRS, stratified by genetic ancestry assignment.

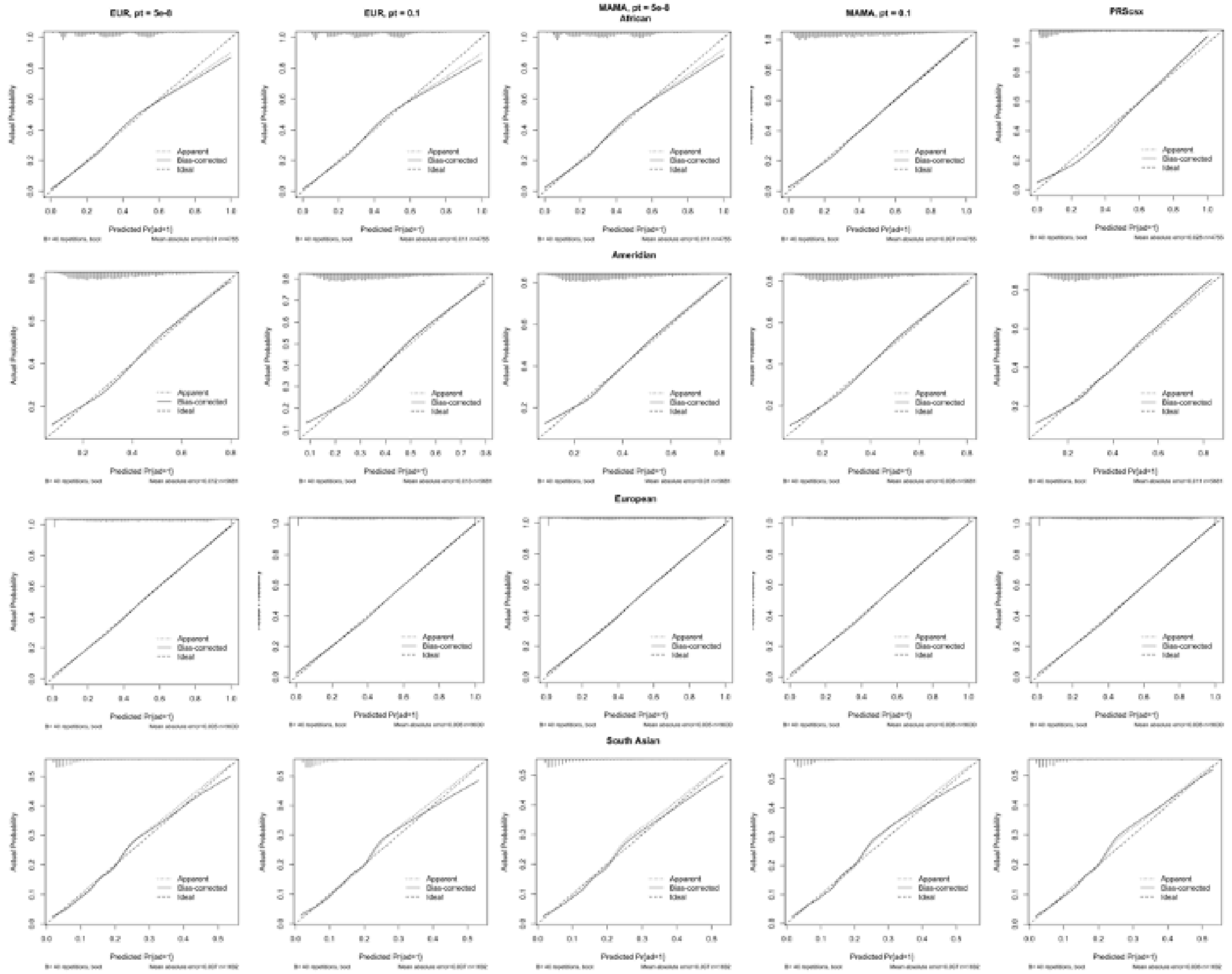


Supplementary figure 5. Calibration pot for PRS across ancestry groups. Calibration plots comparing predicted and observed AD risk for all PRS models across ancestry groups. The x-axis represents predicted disease risk, and the y-axis shows the apparent, bias-corrected and ideal calibration curves.

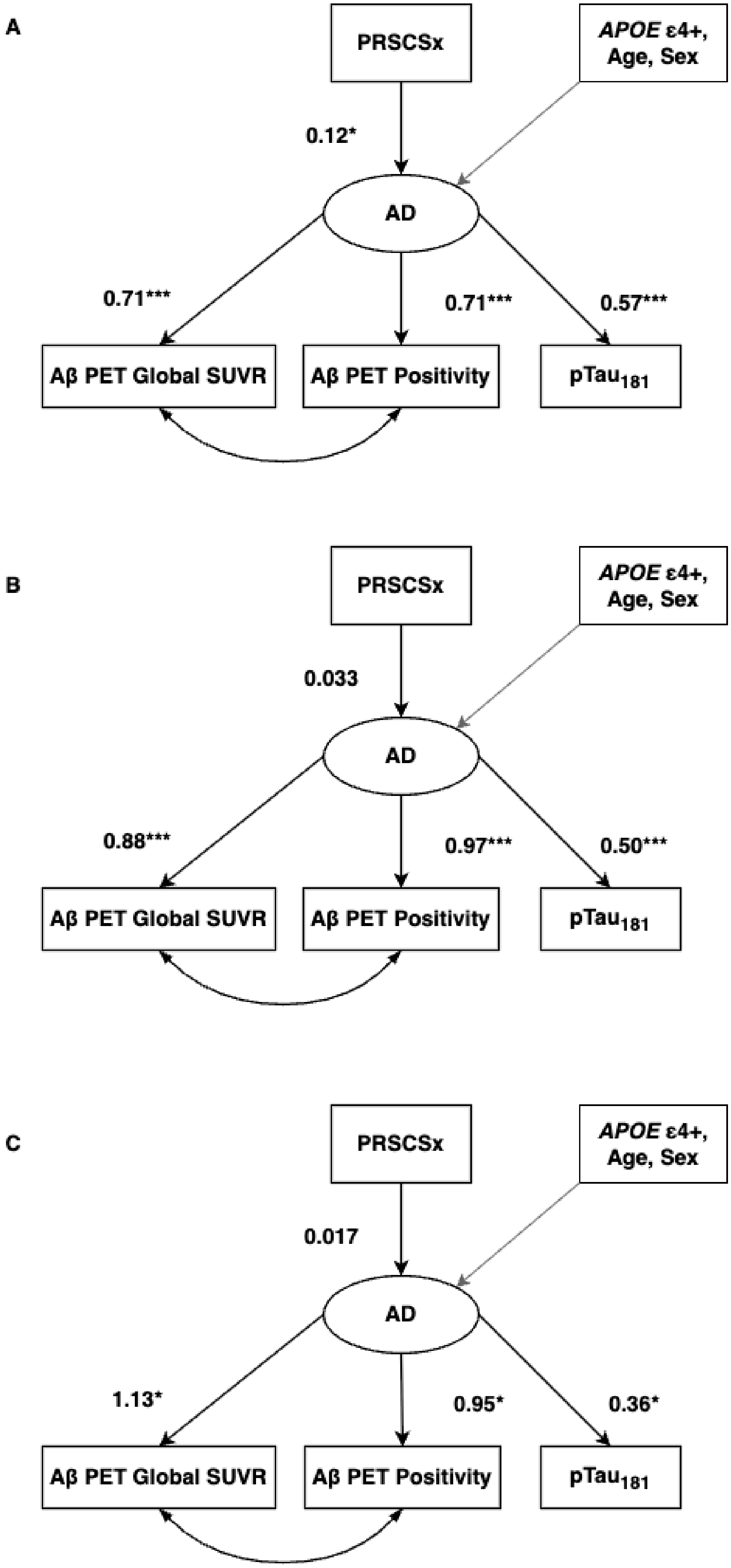

A
PRSCSx
APOE ε4+, Age, Sex
0.12*
AD
0.71***
0.71***
0.57***
Aβ PET Global SUVR
Aβ PET Positivity
pTau181
B
PRSCSx
APOE ε4+, Age, Sex
0.033
AD
0.88***
0.97***
0.50***
Aβ PET Global SUVR
Aβ PET Positivity
pTau181
C
PRSCSx
APOE ε4+, Age, Sex
0.017
AD
1.13*
0.95*
0.36*
Aβ PET Global SUVR
Aβ PET Positivity
pTau181

Supplementary figure 6. Multigroup path analysis in HABSHD for the participants of A) European (N = 1,040), B) Amerindian (N = 890), and C) African ancestry (N = 363). CFI = 0.986, TLI = 0.969, RMSEA = 0.034, SRMR = 0.025
