## Supplementary Tables 1-6 for "Cross-Ancestry Polygenic Risk Scores Enhance Alzheimer’s Disease Risk Prediction in Multiethnic Cohorts"

**Supplementary Table 1. Associations and predictive performance of PRSCSx cross-ancestry PRS model to AD diagnosis in the ADSP. P-values were adjusted for FDR at 5%. African (AFR), Amerindian (AMR), and European (EUR)**

| Ancesty | Ancestry normalized | Model + P-value threshold | Odds Ratio | Lower Conf. Interval | Upper Conf. Interval | P-value | P-value, FDR adj. | R² | AUC |
|---|---|---|---|---|---|---|---|---|---|
| EUR | Base | Base | NA | NA | NA | NA | NA | 0.40 | 0.82 |
| EUR | Sum | EUR, Pt = 5e-8 | 1.08 | 1.03E+00 | 1.14 | 1.97E-03 | 3.29E-03 | 0.41 | 0.82 |
| EUR | Sum | EUR, Pt = 0.1 | 1.10 | 1.03E+00 | 1.18 | 3.04E-03 | 4.59E-03 | 0.41 | 0.82 |
| EUR | Sum | MAMA, Pt = 5e-8 | 1.36 | 1.29E+00 | 1.43 | 1.55E-30 | 2.33E-29 | 0.42 | 0.82 |
| EUR | Sum | MAMA, Pt = 0.1 | 1.36 | 1.28E+00 | 1.43 | 1.03E-25 | 5.18E-25 | 0.42 | 0.82 |
| EUR | Sum | PRSCSx | 1.36 | 1.28E+00 | 1.45 | 3.02E-22 | 1.01E-21 | 0.42 | 0.82 |
| EUR | Normalized | EUR, Pt = 5e-8 | 1.09 | 1.03E+00 | 1.15 | 1.85E-03 | 3.26E-03 | 0.41 | 0.82 |
| EUR | Normalized | EUR, Pt = 0.1 | 1.09 | 1.03 | 1.15 | 3.06E-03 | 4.59E-03 | 0.41 | 0.82 |
| EUR | Normalized | MAMA, Pt = 5e-8 | 1.37 | 1.30 | 1.44 | 1.40E-30 | 2.33E-29 | 0.42 | 0.82 |
| EUR | Normalized | MAMA, Pt = 0.1 | 1.34 | 1.27 | 1.41 | 1.04E-25 | 5.18E-25 | 0.42 | 0.82 |
| EUR | Normalized | PRSCSx | 1.31 | 1.24 | 1.38 | 3.67E-22 | 1.10E-21 | 0.42 | 0.82 |
| AFR | Base | Base | NA | NA | NA | NA | NA | 0.42 | 0.84 |
| AFR | Sum | EUR, Pt = 5e-8 | 0.98 | 8.43E-01 | 1.15 | 0.84 | 0.93 | 0.42 | 0.84 |
| AFR | Sum | EUR, Pt = 0.1 | 0.99 | 0.91 | 1.08 | 0.79 | 0.93 | 0.42 | 0.84 |
| AFR | Sum | MAMA, Pt = 5e-8 | 1.19 | 1.01 | 1.39 | 0.04 | 0.05 | 0.42 | 0.84 |
| AFR | Sum | MAMA, Pt = 0.1 | 2.12 | 1.85 | 2.42 | 1.13E-27 | 1.13E-26 | 0.49 | 0.87 |
| AFR | Sum | PRSCSx | 1.53 | 1.39E+00 | 1.68 | 1.59E-18 | 3.97E-18 | 0.46 | 0.86 |
| AFR | Normalized | EUR, Pt = 5e-8 | 0.99 | 8.68E-01 | 1.12 | 0.83 | 0.93 | 0.42 | 0.84 |
| AFR | Normalized | EUR, Pt = 0.1 | 0.99 | 0.88 | 1.12 | 0.87 | 0.93 | 0.42 | 0.84 |
| AFR | Normalized | MAMA, Pt = 5e-8 | 1.13 | 1.00 | 1.27 | 0.05 | 0.06 | 0.42 | 0.84 |
| AFR | Normalized | MAMA, Pt = 0.1 | 1.80 | 1.62 | 2.01 | 1.95E-27 | 1.46E-26 | 0.49 | 0.87 |
| AFR | Normalized | PRSCSx | 1.71 | 1.52E+00 | 1.93 | 7.27E-19 | 1.98E-18 | 0.46 | 0.86 |
| AMR | Base | Base | NA | NA | NA | NA | NA | 0.45 | 0.86 |
| AMR | Sum | EUR, Pt = 5e-8 | 1 | 9.36E-01 | 1.07 | 0.99 | 0.99 | 0.45 | 0.86 |
| AMR | Sum | EUR, Pt = 0.1 | 0.95 | 0.89 | 1.01 | 0.09 | 0.12 | 0.45 | 0.86 |
| AMR | Sum | MAMA, Pt = 5e-8 | 1.19 | 1.11 | 1.28 | 5.37E-07 | 1.07E-06 | 0.45 | 0.86 |
| AMR | Sum | MAMA, Pt = 0.1 | 1.42 | 1.33E+00 | 1.52 | 2.72E-25 | 1.17E-24 | 0.46 | 0.86 |
| AMR | Sum | PRSCSx | 1.31 | 1.23E+00 | 1.40 | 1.31E-16 | 3.03E-16 | 0.46 | 0.86 |
| AMR | Normalized | EUR, Pt = 5e-8 | 1.00 | 9.39E-01 | 1.07 | 0.97 | 0.99 | 0.45 | 0.86 |
| AMR | Normalized | EUR, Pt = 0.1 | 0.95 | 0.90 | 1.01 | 0.12 | 0.15 | 0.45 | 0.86 |
| AMR | Normalized | MAMA, Pt = 5e-8 | 1.17 | 1.10 | 1.24 | 9.28E-07 | 1.74E-06 | 0.45 | 0.86 |
| AMR | Normalized | MAMA, Pt = 0.1 | 1.34 | 1.26 | 1.41 | 6.83E-24 | 2.56E-23 | 0.46 | 0.86 |
| AMR | Normalized | PRSCSx | 1.29 | 1.21 | 1.37 | 1.06E-15 | 2.26E-15 | 0.46 | 0.86 |

**Supplementary Table 2. Association of PRSCSx cross-ancestry PRS model to AD endophenotypes in the HABSHD cohort, stratified by ancestry. African (AFR), Amerindian (AMR), and European (EUR). P-values were adjusted for FDR at 5%.**

| Ancestry | Odds Ratio | Lower Conf. Interval | Upper Conf. Interval | P-value | P-value, FDR adj. | Category | Outcome |
|---|---|---|---|---|---|---|---|
| AFR | 0.94 | 0.85 | 1.05 | 0.27 | 0.792037037 | Plasma Biomarkers | Aβ40 |
| AFR | 0.94 | 0.85 | 1.04 | 0.20 | 0.747042254 | Plasma Biomarkers | Aβ42 |
| AFR | 0.18 | 0.00 | 55.24 | 0.56 | 0.89746988 | Plasma Biomarkers | Aβ42/Aβ40 |
| AFR | 0.94 | 0.80 | 1.10 | 0.44 | 0.833890909 | Cognitive Function | CDR |
| AFR | 0.99 | 0.98 | 1.00 | 0.15 | 0.684210526 | Neuroimaging | Cortical Thickness Meta-ROI |
| AFR | 0.97 | 0.92 | 1.03 | 0.35 | 0.796595745 | Cognitive Function | Executive Function |
| AFR | 1.06 | 0.98 | 1.14 | 0.14 | 0.668571429 | Neuroimaging | Hippocampal Volume |
| AFR | 0.97 | 0.90 | 1.05 | 0.44 | 0.833890909 | Cognitive Function | Memory |
| AFR | 0.88 | 0.71 | 1.09 | 0.25 | 0.78313253 | Cognitive Function | MMSE |
| AFR | 0.99 | 0.92 | 1.07 | 0.76 | 0.941358314 | Plasma Biomarkers | NfL |
| AFR | 1.00 | 0.91 | 1.09 | 0.96 | 0.980512821 | Plasma Biomarkers | pTau |
| AFR | 0.98 | 0.89 | 1.09 | 0.72 | 0.925728395 | Plasma Biomarkers | Total Tau |
| AFR | 0.98 | 0.90 | 1.06 | 0.57 | 0.89746988 | Cognitive Function | Verbal Ability |
| all | 0.97 | 0.93 | 1.02 | 0.23 | 0.776666667 | Plasma Biomarkers | Aβ40 |
| all | 0.96 | 0.92 | 1.00 | 0.04 | 0.449090909 | Plasma Biomarkers | Aβ42 |
| all | 0.79 | 0.23 | 2.68 | 0.71 | 0.925728395 | Plasma Biomarkers | Aβ42/Aβ40 |
| all | 0.99 | 0.93 | 1.05 | 0.69 | 0.923701799 | Cognitive Function | CDR |
| all | 1.00 | 0.99 | 1.01 | 0.91 | 0.979710145 | Neuroimaging | Cortical Thickness Meta-ROI |
| all | 0.98 | 0.96 | 1.01 | 0.21 | 0.753103448 | Cognitive Function | Executive Function |
| all | 0.98 | 0.95 | 1.02 | 0.42 | 0.832 | Neuroimaging | Hippocampal Volume |
| all | 0.99 | 0.95 | 1.03 | 0.56 | 0.89746988 | Cognitive Function | Memory |
| all | 1.01 | 0.89 | 1.13 | 0.93 | 0.980512821 | Cognitive Function | MMSE |
| all | 1.02 | 0.98 | 1.06 | 0.30 | 0.792037037 | Plasma Biomarkers | NfL |
| all | 1.05 | 1.00 | 1.09 | 0.03 | 0.442 | Plasma Biomarkers | pTau |
| all | 1.02 | 0.97 | 1.06 | 0.50 | 0.882047782 | Plasma Biomarkers | Total Tau |
| all | 1.02 | 0.98 | 1.06 | 0.33 | 0.792037037 | Cognitive Function | Verbal Ability |
| AMR | 0.95 | 0.89 | 1.02 | 0.20 | 0.747042254 | Plasma Biomarkers | Aβ40 |
| AMR | 0.96 | 0.89 | 1.04 | 0.29 | 0.792037037 | Plasma Biomarkers | Aβ42 |
| AMR | 1.64 | 0.39 | 6.91 | 0.50 | 0.886644068 | Plasma Biomarkers | Aβ42/Aβ40 |
| AMR | 1.03 | 0.91 | 1.16 | 0.65 | 0.915862069 | Cognitive Function | CDR |
| AMR | 1.01 | 1.00 | 1.02 | 0.25 | 0.78313253 | Neuroimaging | Cortical Thickness Meta-ROI |
| AMR | 0.98 | 0.93 | 1.03 | 0.35 | 0.796595745 | Cognitive Function | Executive Function |
| AMR | 0.94 | 0.88 | 1.00 | 0.05 | 0.458305085 | Neuroimaging | Hippocampal Volume |

| | | | | | | | |
|---|---|---|---|---|---|---|---|
| AMR | 1.02 | 0.96 | 1.09 | 0.47 | 0.859368421 | Cognitive Function | Memory |
| AMR | 1.14 | 0.88 | 1.47 | 0.33 | 0.792037037 | Cognitive Function | MMSE |
| AMR | 1.02 | 0.95 | 1.09 | 0.61 | 0.911494253 | Plasma Biomarkers | NfL |
| AMR | 1.01 | 0.94 | 1.09 | 0.75 | 0.939759036 | Plasma Biomarkers | pTau |
| AMR | 1.01 | 0.93 | 1.08 | 0.90 | 0.977991632 | Plasma Biomarkers | Total Tau |
| AMR | 1.03 | 0.97 | 1.11 | 0.32 | 0.792037037 | Cognitive Function | Verbal Ability |
| EUR | 1.01 | 0.95 | 1.08 | 0.71 | 0.925728395 | Plasma Biomarkers | Aβ40 |
| EUR | 0.96 | 0.91 | 1.03 | 0.24 | 0.778343949 | Plasma Biomarkers | Aβ42 |
| EUR | 0.77 | 0.29 | 2.03 | 0.59 | 0.901231672 | Plasma Biomarkers | Aβ42/Aβ40 |
| EUR | 0.98 | 0.91 | 1.06 | 0.68 | 0.919164491 | Cognitive Function | CDR |
| EUR | 1.00 | 0.99 | 1.01 | 0.94 | 0.980512821 | Neuroimaging | Cortical Thickness Meta-ROI |
| EUR | 0.99 | 0.96 | 1.03 | 0.65 | 0.915862069 | Cognitive Function | Executive Function |
| EUR | 0.98 | 0.93 | 1.05 | 0.60 | 0.906976744 | Neuroimaging | Hippocampal Volume |
| EUR | 0.97 | 0.92 | 1.02 | 0.23 | 0.776578947 | Cognitive Function | Memory |
| EUR | 0.99 | 0.87 | 1.13 | 0.89 | 0.977468354 | Cognitive Function | MMSE |
| EUR | 1.03 | 0.98 | 1.10 | 0.26 | 0.790643275 | Plasma Biomarkers | NfL |
| EUR | 1.08 | 1.01 | 1.15 | 0.02 | 0.334285714 | Plasma Biomarkers | pTau |
| EUR | 1.03 | 0.96 | 1.11 | 0.38 | 0.816842105 | Plasma Biomarkers | Total Tau |
| EUR | 1.03 | 0.97 | 1.09 | 0.36 | 0.796595745 | Cognitive Function | Verbal Ability |

**Supplementary Table 3. Association of PRSCSx cross-ancestry PRS model to AD endophenotypes in the ADSP cohort, total. P-values were adjusted for FDR at 5%. MASS::polr package was used to run ordinal regression for Braak staging, Thal phase, and CERAD outcomes.**

| Ancestry | Variable | Category | Estimates | Std.Err | Lower Conf. Interval | Upper Conf. Interval | $R^2$ | P-value | P-value, FDR adj. |
|---|---|---|---|---|---|---|---|---|---|
| all | Aβ42 | CSF Biomarkers | -0.091 | 0.034 | -0.16 | -0.023 | 0.17 | 0.0084 | 0.015 |
| all | pTau | CSF Biomarkers | 0.052 | 0.035 | -0.017 | 0.12 | 0.084 | 0.14 | 0.14 |
| all | Tau | CSF Biomarkers | 0.054 | 0.035 | -0.016 | 0.12 | 0.072 | 0.13 | 0.14 |
| all | BRAAK | Neuroimaging | 1.09 | 0.035 | 1.03 | 1.16 | 0.11 | 0.011 | 0.016 |
| all | Thal Phase | Neuroimaging | 1.10 | 0.059 | 0.98 | 1.21 | 0.11 | 0.115 | 0.14 |
| all | CERAD | Neuroimaging | 1.12 | 0.038 | 1.05 | 1.20 | 0.11 | 0.0022 | 0.0050 |
| all | Memory | Cognitive | -0.065 | 0.0073 | -0.079 | -0.051 | 0.25 | 5.34E-19 | 9.62E-18 |
| all | Language | Cognitive | -0.051 | 0.0070 | -0.065 | -0.038 | 0.22 | 1.84E-13 | 1.10E-12 |
| all | Executive Function | Cognitive | -0.049 | 0.0075 | -0.064 | -0.034 | 0.30 | 8.68E-11 | 3.13E-10 |
| all | Aβ42 | CSF Biomarkers | -0.084 | 0.034 | -0.15 | -0.017 | 0.17 | 0.015 | 0.026 |
| all | pTau | CSF Biomarkers | 0.035 | 0.036 | -0.036 | 0.11 | 0.077 | 0.33 | 0.35 |
| all | Tau | CSF Biomarkers | 0.049 | 0.035 | -0.021 | 0.12 | 0.070 | 0.17 | 0.23 |
| AFR | Memory | Cognitive | -0.063 | 0.027 | -0.12 | -0.0093 | 0.36 | 0.022 | 0.036 |
| AFR | Executive Function | Cognitive | -0.033 | 0.034 | -0.10 | 0.034 | 0.38 | 0.33 | 0.35 |
| AFR | Language | Cognitive | -0.0094 | 0.028 | -0.064 | 0.045 | 0.40 | 0.737 | 0.74 |
| AMR | Language | Cognitive | -0.026 | 0.010 | -0.047 | -0.0058 | 0.29 | 0.012 | 0.024 |
| AMR | Executive Function | Cognitive | -0.015 | 0.012 | -0.039 | 0.0091 | 0.34 | 0.22 | 0.29 |
| AMR | Memory | Cognitive | -0.014 | 0.012 | -0.036 | 0.0091 | 0.32 | 0.24 | 0.29 |
| EUR | Memory | Cognitive | -0.080 | 0.0092 | -0.099 | -0.062 | 0.26 | 4.13E-18 | 3.71E-17 |
| EUR | Language | Cognitive | -0.059 | 0.0091 | -0.077 | -0.042 | 0.11 | 6.37E-11 | 2.87E-10 |
| EUR | Executive Function | Cognitive | -0.056 | 0.010 | -0.075 | -0.037 | 0.19 | 4.05E-09 | 1.22E-08 |

**Supplementary Table 4. Association of PRSCSx cross-ancestry PRS model to AD latent variable in the HABSHD cohort, total.**

| Metric | Value |
|---|---|
| Estimator | FIML |
| Optimization method | NLMINB |
| Number of model parameters | 17 |
| Number of observations (Used) | 2029 |
| Number of observations (Total) | 2559 |
| Number of missing patterns | 4 |
| User Model Test statistic | 21.859 |
| User Model Degrees of freedom | 13 |
| User Model P-value (Chi-square) | 0.058 |
| Baseline Model Test statistic | 1144.85 |
| Baseline Model Degrees of freedom | 24 |
| Baseline Model P-value | 0 |
| Comparative Fit Index (CFI) | 0.992 |
| Tucker-Lewis Index (TLI) | 0.985 |
| Robust Comparative Fit Index (CFI) | 0.992 |
| Robust Tucker-Lewis Index (TLI) | 0.986 |
| Loglikelihood user model (H0) | -2091.372 |
| Loglikelihood unrestricted model (H1) | -2080.442 |
| Akaike (AIC) | 4216.743 |
| Bayesian (BIC) | 4312.204 |
| Sample-size adjusted BIC (SABIC) | 4258.193 |
| RMSEA | 0.018 |
| RMSEA 90% CI lower | 0 |
| RMSEA 90% CI upper | 0.031 |
| P-value H0: RMSEA <= 0.050 | 1 |
| P-value H0: RMSEA >= 0.080 | 0 |
| Robust RMSEA | 0.03 |
| Robust RMSEA 90% CI lower | 0 |
| Robust RMSEA 90% CI upper | 0.053 |
| P-value H0: Robust RMSEA <= 0.050 | 0.923 |
| P-value H0: Robust RMSEA >= 0.080 | 0 |
| SRMR | 0.018 |

| Parameter | Estimate | Std.Err | z-value | P(>\|z\|) | Std.lv | Std.all |
|---|---|---|---|---|---|---|
| AD_pathology =~ ptau_residual | 1 | | | | 0.51 | 0.54 |
| AD_pathology =~ ab_pet | 0.56 | 0.062 | 9 | < 0.001 | 0.28 | 0.72 |
| AD_pathology =~ ab_suvr | 0.25 | 0.026 | 9.44 | < 0.001 | 0.13 | 0.78 |
| AD_pathology ~ prs_z_prscsx | 0.042 | 0.017 | 2.47 | 0.014 | 0.083 | 0.082 |
| AD_pathology ~ cogd | 0.18 | 0.019 | 9.77 | < 0.001 | 0.36 | 0.36 |
| AD_pathology ~ apoe4 | 0.37 | 0.041 | 9.03 | < 0.001 | 0.73 | 0.33 |
| AD_pathology ~ PC1 | -0.0020 | 0.0060 | -0.33 | 0.74 | -0.0040 | -0.12 |
| AD_pathology ~ PC2 | 0.0050 | 0.010 | 0.46 | 0.65 | 0.0090 | 0.21 |
| AD_pathology ~ PC3 | 0.0040 | 0.010 | 0.42 | 0.67 | 0.0080 | 0.17 |
| AD_pathology ~ PC4 | -0.018 | 0.014 | -1.28 | 0.20 | -0.035 | -0.42 |
| .ab_pet ~~ .ab_suvr | 0.016 | 0.004 | 4.08 | < 0.001 | 0.016 | 0.59 |
| .ptau_residual intercept | -0.18 | 0.35 | -0.53 | 0.60 | -0.18 | -0.20 |
| .ab_pet intercept | 0.12 | 0.19 | 0.60 | 0.55 | 0.12 | 0.30 |
| .ab_suvr intercept | 1.01 | 0.086 | 11.81 | < 0.001 | 1.01 | 6.30 |
| .ptau_residual variance | 0.62 | 0.034 | 18.55 | < 0.001 | 0.62 | 0.71 |
| .ab_pet variance | 0.076 | 0.010 | 7.37 | < 0.001 | 0.076 | 0.49 |
| .ab_suvr variance | 0.0100 | 0.0020 | 5.67 | < 0.001 | 0.010 | 0.40 |
| AD_pathology variance | 0.19 | 0.027 | 6.90 | < 0.001 | 0.74 | 0.74 |

**Supplementary Table 5. Association of PRSCSx cross-ancestry PRS model to AD latent variable in the HABSHD cohort, stratified by genetic ancestry. African (AFR), Amerindian (AMR), and Europ**

| Metric | Value |
|---|---|
| Estimator | FIML |
| Optimization method | NLMINB |
| Number of model parameters | 51 |
| Number of observations (Used) AFR | 330 |
| Number of observations (Total) AFR | 574 |
| Number of observations (Used) EUR | 995 |
| Number of observations (Total) EUR | 1040 |
| Number of observations (Used) AMR | 704 |
| Number of observations (Total) AMR | 891 |
| Number of missing patterns AFR | 4 |
| Number of missing patterns EUR | 4 |
| Number of missing patterns AMR | 4 |
| User Model Test statistic | 37.499 |
| User Model Degrees of freedom | 39 |
| User Model P-value (Chi-square) | 0.538 |
| User Model Test statistic AFR | 11.102 |
| User Model Test statistic EUR | 8.036 |
| User Model Test statistic AMR | 18.362 |
| Baseline Model Test statistic | 1159.249 |
| Baseline Model Degrees of freedom | 72 |
| Baseline Model P-value | 0 |
| Comparative Fit Index (CFI) | 1 |
| Tucker-Lewis Index (TLI) | 1.003 |
| Robust Comparative Fit Index (CFI) | 0.999 |
| Robust Tucker-Lewis Index (TLI) | 0.998 |
| Loglikelihood user model (H0) | -1994.769 |
| Loglikelihood unrestricted model (H1) | -1976.019 |
| Akaike (AIC) | 4091.537 |
| Bayesian (BIC) | 4377.917 |
| Sample-size adjusted BIC (SABIC) | 4215.887 |
| RMSEA | 0 |
| RMSEA 90% CI lower | 0 |
| RMSEA 90% CI upper | 0.025 |
| P-value H0: RMSEA <= 0.050 | 1 |
| P-value H0: RMSEA >= 0.080 | 0 |
| Robust RMSEA | 0.012 |
| Robust RMSEA 90% CI lower | 0 |
| Robust RMSEA 90% CI upper | 0.048 |
| P-value H0: Robust RMSEA <= 0.050 | 0.959 |
| P-value H0: Robust RMSEA >= 0.080 | 0 |
| SRMR | 0.016 |

| Parameter | Estimate | Std.Err | z-value | P(>\|z\|) | Std.lv | Std.all |
|---|---|---|---|---|---|---|
| # Group: AFR | | | | | | |
| AD_pathology =~ ptau_residual | 1 | | | | 0.38 | 0.42 |
| AD_pathology =~ ab_pet | 0.75 | 0.25 | 2.97 | 0.0030 | 0.29 | 0.80 |
| AD_pathology =~ ab_suvr | 0.31 | 0.10 | 3.07 | 0.0020 | 0.12 | 0.96 |
| AD_pathology ~ prs_z_prscsx | 0.0100 | 0.020 | 0.49 | 0.62 | 0.025 | 0.027 |
| AD_pathology ~ cogd | 0.14 | 0.051 | 2.64 | 0.0080 | 0.35 | 0.29 |
| AD_pathology ~ apoe4 | 0.21 | 0.078 | 2.71 | 0.0070 | 0.55 | 0.27 |
| AD_pathology ~ PC1 | 0.013 | 0.011 | 1.19 | 0.24 | 0.034 | 0.26 |
| AD_pathology ~ PC2 | -0.024 | 0.018 | -1.32 | 0.19 | -0.063 | -0.31 |
| AD_pathology ~ PC3 | 0.026 | 0.019 | 1.34 | 0.18 | 0.068 | 0.095 |
| AD_pathology ~ PC4 | 0.012 | 0.021 | 0.56 | 0.58 | 0.031 | 0.056 |
| .ab_pet ~~ .ab_suvr | 0.0010 | 0.011 | 0.14 | 0.89 | 0.0010 | 0.20 |
| .ptau_residual intercept | -0.95 | 0.62 | -1.52 | 0.13 | -0.95 | -1.03 |
| .ab_pet intercept | -0.39 | 0.43 | -0.91 | 0.36 | -0.39 | -1.09 |
| .ab_suvr intercept | 0.81 | 0.18 | 4.57 | < 0.001 | 0.81 | 6.59 |
| .ptau_residual variance | 0.70 | 0.071 | 9.85 | < 0.001 | 0.70 | 0.83 |
| .ab_pet variance | 0.045 | 0.028 | 1.61 | 0.107 | 0.045 | 0.35 |
| .ab_suvr variance | 0.0010 | 0.004 | 0.27 | 0.79 | 0.0010 | 0.077 |
| AD_pathology variance | 0.13 | 0.046 | 2.74 | 0.006 | 0.86 | 0.86 |
| # Group: EUR | | | | | | |
| AD_pathology =~ ptau_residual | 1.00 | | | | 0.55 | 0.57 |
| AD_pathology =~ ab_pet | 0.57 | 0.089 | 6.40 | < 0.001 | 0.32 | 0.70 |
| AD_pathology =~ ab_suvr | 0.28 | 0.041 | 6.85 | < 0.001 | 0.15 | 0.74 |
| AD_pathology ~ prs_z_prscsx | 0.066 | 0.028 | 2.38 | 0.017 | 0.12 | 0.11 |
| AD_pathology ~ cogd | 0.24 | 0.028 | 8.72 | < 0.001 | 0.44 | 0.45 |
| AD_pathology ~ apoe4 | 0.51 | 0.061 | 8.35 | < 0.001 | 0.92 | 0.42 |
| AD_pathology ~ PC1 | -0.0040 | 0.014 | -0.28 | 0.78 | -0.007 | -0.015 |
| AD_pathology ~ PC2 | 0.014 | 0.019 | 0.72 | 0.47 | 0.025 | 0.044 |
| AD_pathology ~ PC3 | -0.015 | 0.021 | -0.68 | 0.49 | -0.026 | -0.032 |
| AD_pathology ~ PC4 | -0.042 | 0.022 | -1.93 | 0.054 | -0.077 | -0.10 |
| .ab_pet ~~ .ab_suvr | 0.030 | 0.0080 | 3.79 | < 0.001 | 0.030 | 0.65 |
| .ptau_residual intercept | -0.26 | 0.73 | -0.36 | 0.72 | -0.26 | -0.27 |
| .ab_pet intercept | 0.057 | 0.42 | 0.14 | 0.89 | 0.057 | 0.13 |
| .ab_suvr intercept | 0.98 | 0.20 | 4.82 | < 0.001 | 0.98 | 4.69 |
| .ptau_residual variance | 0.64 | 0.054 | 12.02 | < 0.001 | 0.64 | 0.68 |
| .ab_pet variance | 0.10 | 0.018 | 5.71 | < 0.001 | 0.10 | 0.51 |
| .ab_suvr variance | 0.020 | 0.004 | 5.00 | < 0.001 | 0.020 | 0.46 |
| AD_pathology variance | 0.19 | 0.045 | 4.17 | < 0.001 | 0.63 | 0.63 |
| # Group: AMR | | | | | | |
| AD_pathology =~ ptau_residual | 1 | | | | 0.40 | 0.46 |
| AD_pathology =~ ab_pet | 0.82 | 0.24 | 3.44 | < 0.001 | 0.33 | 0.95 |

| | | | | | | | | | |
|---|---|---|---|---|---|---|---|---|---|
| | | | AD_pathology =~ ab_suvr | 0.28 | 0.085 | 3.25 | < 0.001 | 0.11 | 0.92 |
| | | | AD_pathology ~ prs_z_prscsx | 0.025 | 0.029 | 0.85 | 0.40 | 0.062 | 0.053 |
| | | | AD_pathology ~ cogd | 0.10 | 0.029 | 3.39 | < 0.001 | 0.25 | 0.26 |
| | | | AD_pathology ~ apoe4 | 0.22 | 0.075 | 2.98 | 0.0030 | 0.56 | 0.21 |
| | | | AD_pathology ~ PC1 | -0.019 | 0.012 | -1.51 | 0.13 | -0.046 | -0.17 |
| | | | AD_pathology ~ PC2 | 0.0010 | 0.015 | 0.064 | 0.95 | 0.0020 | 0.025 |
| | | | AD_pathology ~ PC3 | 0 | 0.016 | -0.0060 | 1.00 | 0 | -0.0030 |
| | | | AD_pathology ~ PC4 | -0.011 | 0.025 | -0.46 | 0.65 | -0.028 | -0.21 |
| | | | .ab_pet ~~ .ab_suvr | -0.002 | 0.011 | -0.19 | 0.85 | -0.002 | -0.43 |
| | | | .ptau_residual intercept | -0.34 | 0.56 | -0.61 | 0.54 | -0.34 | -0.40 |
| | | | .ab_pet intercept | -0.12 | 0.47 | -0.24 | 0.81 | -0.12 | -0.34 |
| | | | .ab_suvr intercept | 0.94 | 0.16 | 5.87 | < 0.001 | 0.94 | 7.84 |
| | | | .ptau_residual variance | 0.59 | 0.058 | 10.23 | < 0.001 | 0.59 | 0.79 |
| | | | .ab_pet variance | 0.011 | 0.034 | 0.34 | 0.74 | 0.011 | 0.095 |
| | | | .ab_suvr variance | 0.002 | 0.0040 | 0.53 | 0.59 | 0.0020 | 0.15 |
| | | | AD_pathology variance | 0.14 | 0.047 | 2.93 | 0.0030 | 0.86 | 0.86 |

**Supplementary Table 6. Comparison of $R^2$ and differences ($\Delta R^2$) between single-ancestry, multi-ancestry (MAMA), and cross-ancestry (PRS-CSx) PRS models across African (AFR), Amerindian (AMR), and European (EUR) ancestry groups. $R^2$ estimates, standard errors (SE), confidence intervals, and p-values were derived using the $\Delta R^2$ framework described in Momin et al. (2023, AJHG).**

| Ancestry | PRS1 | PRS2 | $R^2$, PRS1 | $R^2$, PRS2 | $\Delta R^2$ | Lower Conf. Interval | Upper Conf. Interval | P-value | SE, PRS1 | SE, PRS2 | ΔSE | Comparison |
|---|---|---|---|---|---|---|---|---|---|---|---|---|
| AFR | MAMA, GWS | EUR, GWS | 2.59E-03 | 9.03E-04 | 1.69E-03 | -2.01E-03 | 5.38E-03 | 0.37 | 2.38E-03 | 1.52E-03 | 1.88E-03 | MAMA, GWS x EUR, GWS |
| AFR | PRSCSx | EUR, GWS | 0.047 | 9.03E-04 | 0.046 | 0.028 | 0.064 | 4.65E-07 | 9.28E-03 | 1.52E-03 | 9.19E-03 | PRSCSx x EUR, GWS |
| AFR | PRSCSx | MAMA, GWS | 0.047 | 2.59E-03 | 0.045 | 0.027 | 0.063 | 1.30E-06 | 9.28E-03 | 2.38E-03 | 9.22E-03 | PRSCSx x MAMA, GWS |
| AFR | MAMA, 0.1 | EUR, 0.1 | 0.076 | 5.31E-04 | 0.075 | 0.052 | 0.098 | 1.83E-10 | 0.011 | 1.25E-03 | 0.012 | MAMA, 0.1 x EUR, 0.1 |
| AFR | PRSCSx | EUR, 0.1 | 0.047 | 5.31E-04 | 0.047 | 0.028 | 0.065 | 8.07E-07 | 9.28E-03 | 1.25E-03 | 9.46E-03 | PRSCSx x EUR, 0.1 |
| AFR | PRSCSx | MAMA, 0.1 | 0.047 | 0.076 | -0.028 | -0.051 | -5.73E-03 | 0.014 | 9.28E-03 | 1.14E-02 | 0.012 | PRSCSx x MAMA, 0.1 |
| AMR | MAMA, GWS | EUR, GWS | 4.55E-03 | 4.18E-03 | 3.71E-04 | -2.10E-03 | 2.84E-03 | 0.77 | 1.61E-03 | 1.55E-03 | 1.26E-03 | MAMA, GWS x EUR, GWS |
| AMR | PRSCSx | EUR, GWS | 0.010 | 4.18E-03 | 6.03E-03 | 1.13E-03 | 0.011 | 0.016 | 2.39E-03 | 1.55E-03 | 2.50E-03 | PRSCSx x EUR, GWS |
| AMR | PRSCSx | MAMA, GWS | 0.010 | 4.55E-03 | 5.66E-03 | 7.91E-04 | 0.011 | 0.023 | 2.39E-03 | 1.61E-03 | 2.48E-03 | PRSCSx x MAMA, GWS |
| AMR | MAMA, 0.1 | EUR, 0.1 | 0.018 | 2.19E-03 | 0.016 | 0.011 | 0.022 | 6.36E-09 | 3.17E-03 | 1.13E-03 | 2.78E-03 | MAMA, 0.1 x EUR, 0.1 |
| AMR | PRSCSx | EUR, 0.1 | 0.010 | 2.19E-03 | 8.03E-03 | 3.36E-03 | 0.013 | 7.54E-04 | 2.39E-03 | 1.13E-03 | 2.38E-03 | PRSCSx x EUR, 0.1 |
| AMR | PRSCSx | MAMA, 0.1 | 0.010 | 0.018 | -8.14E-03 | -0.014 | -1.78E-03 | 0.012 | 2.39E-03 | 3.17E-03 | 3.24E-03 | PRSCSx x MAMA, 0.1 |
| EUR | MAMA, GWS | EUR, GWS | 0.018 | 0.026 | -8.48E-03 | -0.013 | -3.97E-03 | 2.27E-04 | 2.84E-03 | 3.43E-03 | 2.30E-03 | MAMA, GWS x EUR, GWS |
| EUR | PRSCSx | EUR, GWS | 0.013 | 0.026 | -0.013 | -0.020 | -5.94E-03 | 2.62E-04 | 2.48E-03 | 3.43E-03 | 3.52E-03 | PRSCSx x EUR, GWS |
| EUR | PRSCSx | MAMA, GWS | 0.013 | 0.018 | -4.36E-03 | -0.010 | 1.72E-03 | 0.16 | 2.48E-03 | 2.84E-03 | 3.10E-03 | PRSCSx x MAMA, GWS |
| EUR | MAMA, 0.1 | EUR, 0.1 | 0.015 | 0.018 | -2.24E-03 | -6.96E-03 | 2.49E-03 | 0.35 | 2.67E-03 | 2.85E-03 | 2.41E-03 | MAMA, 0.1 x EUR, 0.1 |
| EUR | PRSCSx | EUR, 0.1 | 0.013 | 0.018 | -4.49E-03 | -0.011 | 1.70E-03 | 0.15 | 2.48E-03 | 2.85E-03 | 3.16E-03 | PRSCSx x EUR, 0.1 |
| EUR | PRSCSx | MAMA, 0.1 | 0.013 | 0.015 | -2.26E-03 | -7.96E-03 | 3.45E-03 | 0.44 | 2.48E-03 | 2.67E-03 | 2.91E-03 | PRSCSx x MAMA, 0.1 |
